# Associations between everyday activities and arterial spin labelling-derived cerebral blood flow: A longitudinal study in community-dwelling elderly volunteers

**DOI:** 10.1101/2022.09.22.22280237

**Authors:** Anne-Marthe Sanders, Geneviève Richard, Knut Kolskår, Kristine M. Ulrichsen, Dag Alnæs, Dani Beck, Erlend S. Dørum, Andreas Engvig, Martina Jonette Lund, Wibeke Nordhøy, Mads L. Pedersen, Jaroslav Rokicki, Jan Egil Nordvik, Lars T. Westlye

## Abstract

**Background:** Cerebral blood flow (CBF) is critical for brain metabolism and overall function. Age-related changes in CBF have been associated with cognitive deficits and increased risk of neurocognitive disorders and vascular events such as stroke. Exercise is considered among the protective factors for age-related brain and cognitive impairment, but how different lifestyle characteristics, such as low or moderate-to-vigorous physical activity or frequency of everyday activities, relate to cross-sectional and longitudinal measures of CBF has not been established.

**Objective:** With the main aim of identifying potential targets for interventions, our objective was to examine associations between cortical and subcortical CBF and frequency of diverse everyday activities in healthy community-dwelling adults aged 65-89 years, and assess to which degree activity level at baseline is associated with longitudinal changes in CBF across a one-to-two years interval.

**Method:** One hundred nineteen (*N* = 119) adults underwent brain magnetic resonance imaging (MRI), neurocognitive, physical, and activity assessment at baseline. CBF was obtained using pseudo-continuous arterial spin labelling (ASL) MRI. Frequency of everyday activities were measured using Frenchay Activities index, while minutes of low and moderate-to-vigorous intensity physical activity were measured using a StepWatch Activity Monitor. Eighty-six participants completed a follow-up ASL MRI, on average 506 (SD = 113) days after the baseline scan.

**Results:** Bayesian multilevel modelling revealed positive associations between baseline cortical and subcortical CBF and various everyday activities. Higher baseline accumbens, putamen and pallidum CBF was associated with more time spent on low intensity (> 0 steps/minute to < 100 steps/minute) physical activity, higher accumbens and caudate CBF with more moderate to vigorous intensity (≥ 100 steps/minute) physical activity, higher cerebral cortical CBF with more participation in social activity, higher cortical and thalamic CBF with more reading, and higher baseline pallidum CBF with more actively pursuing hobbies. We did not find evidence for an association between baseline activity level and longitudinal changes in CBF.

**Conclusion:** The identified associations between everyday activity measures and CBF provide new knowledge on malleable lifestyle factors that may indicate or contribute to healthy brain aging. In addition to the relevance for prioritizing targets for public health guidelines, our findings contribute to disclose parts of the intricate connection between brain metabolism and everyday activities in aging.

## 1. Introduction

The world population aged 60 years and older is rapidly increasing and will account for 22% of the total population by 2050, up from 12% in 2015 (World Health Organization, 2018). Aging increases the risk of a multitude of conditions, including dementia and cardiovascular diseases, with potential severe impact on cognitive functions (Jaul & Barron, 2017) and independence (Carpenter et al., 2006; Sturm et al., 2002). Providing empirical public health advice and interventions aimed at delaying or preventing age-related diseases are important visions of the clinical neurosciences (Gorelick et al., 2011). Lifestyle factors such as physical activity and social participation are gaining increasing attention as indicators of successful cognitive aging and candidate targets for interventions (Chiuve et al., 2008; Forbes et al., 2013; Haeger et al., 2019; Hersi et al., 2017; Sarikaya et al., 2015).

A healthy vascular system is critical for sustaining the substantial metabolic demands of the brain (Attwell et al., 2010). Age-related cerebrovascular changes include arterial stiffness and thickening (Holland et al., 2017), arteriosclerosis (Wang & Bennett, 2012) and reduced cerebral blood flow (CBF) (Alisch et al., 2021), which represent risk factors for cognitive impairments (Brown & Thore, 2011), stroke (Bangen et al., 2009) and dementia in older adults (Gorelick et al., 2011; Kelleher & Soiza, 2013). A better understanding of the associations between amendable lifestyle factors and cerebrovascular function can contribute to the development of targeted interventions and public health advice (Joris et al., 2018).

CBF can be measured non-invasively using ASL sequences (Detre & Alsop, 1999; Williams et al., 1992) in MRI. ASL has revealed decreased cortical and subcortical CBF in older adults (Alisch et al., 2021; Chen et al., 2011; Stoquart-ElSankari et al., 2007; Zhang et al., 2017), which has been associated with everyday activity measures such as regular exercise, fitness or intensity of daily physical activity, albeit with inconclusive results. An intervention study including patients with Alzheimer’s disease (AD) showed no effect of moderate-to-vigorous aerobic training for 16 weeks on global or regional CBF (van der Kleij et al., 2018). Studies on healthy participants have suggested that cardiovascular fitness (Dougherty et al., 2020; Zimmerman et al., 2014) and aerobic training (Kleinloog et al., 2019) mitigate age-related reduction in CBF. However, only one previous cross-sectional study including 52 cognitive healthy adults aged 65-82 years has shown an association between accelerometer-measured physical activity, including sedentary behavior, and CBF in frontal and medial temporal regions (Zlatar et al., 2019). The findings provide correlational support to the hypothesis that more regular activity, and not only exercise, can serve as an indicator for, or even delay, age-related neurovascular changes. Previous studies have also demonstrated beneficial effects of exercise (Hillman et al., 2008; Kramer & Colcombe, 2018), higher cardiorespiratory fitness level (Sokołowski et al., 2021) and participation in everyday activities (Chan et al., 2018; Fratiglioni et al., 2004; Gow et al., 2017) on various cognitive functions in older adults. Importantly, the evidence of beneficial effects of high intensity exercise remains unclear, and a recent 5-year randomized controlled trial revealed no additional effects of exercise with higher intensity compared to following national physical activity guidelines on brain volume (Pani et al., 2021) or cognition (Sokołowski et al., 2021) after the age of 70.

To date, there is a lack of cross-sectional and longitudinal studies investigating the association between late life CBF and engagement in specific everyday activities, encompassing housework, work, social participation and various leisure activities, and few studies have investigated the association between late life CBF and objectively measured physical activity level. The joint and unique associations between the brain vascular system and everyday activities are of particular interest due to the possibilities of low-threshold interventions aiming at promoting brain health in the aging population.

With the objective to test for associations between baseline activity level and CBF at baseline and follow-up, 119 community-dwelling adults aged 65-89 years were assessed using multimodal brain MRI and accelerometer measured minutes of physical activity and completed questionnaires addressing level of different activities, including home maintenance, housework, work, social activities, and various leisure activities. Follow-up MRI (*n*= 86) was performed on average 506 (SD = 113) days after the initial assessment, allowing for longitudinal CBF assessment. Based on previous work indicating lower cortical and subcortical CBF as a risk-factor for age related diseases (Bangen et al., 2009; Kelleher & Soiza, 2013) and sensitivity to lifestyle activity level (Zlatar et al., 2019), we hypothesized that participants with higher activity level at baseline would show higher cortical and subcortical CBF at both baseline and follow-up compared to participants with lower activity level. Further, based on its putative protective factor we hypothesized that overall higher activity level at baseline would be associated with reduced longitudinal decline in cortical and subcortical CBF.

## 2. Materials and methods

### 2.1 Participants

The initial sample consisted of 341 participants from the StrokeMRI project, which aims to investigate lifestyle predictors of brain and cognitive health, including ageing and stroke. Enrolment criteria and recruitment procedures have previously been described (Richard et al., 2018; Sanders et al., 2021). In brief, participants were healthy volunteers aged 18-94 years at enrolment, without history of neurological or psychiatric diseases, or current drug and/or alcohol abuse, and no MRI contraindications (e.g., pacemaker, ferrous implants, pregnancy, claustrophobia). Among the total sample, 131 participants aged 65-89 years underwent an extended protocol including different measures of activity level. Five participants did not complete the MRI ASL protocol, five were excluded due to insufficient MRI data quality, one did not complete the measures of activity level, and one was identified as an outlier and potential influential case on all baseline analysis, and excluded from the analysis.

Accordingly, we included 119 participants at baseline. All individuals were considered to have normal cognitive function at baseline, with a mini-mental state exam (MMSE) score above 24 (Folstein et al., 1975). A follow-up MRI scan was completed, on average, 506 (SD = 113) days after the baseline scan, including 86 of the 119 participants. The Regional Committees for Medical and Health Research Ethics for the South-Eastern Norway approved the study (REK approvals 2014/ 694, 2015/1282). Each participant gave written informed consent at enrolment, in agreement with the Declaration of Helsinki.

### 2.2 Measures of frequency of lifestyle activities

To measure the frequency of engagement in general activities, subscores from the standardized Frenchay Activities Index (FAI) questionnaire (Holbrook & Skilbeck, 1983) was applied. FAI originally consists of a 15-item scale, capturing the regularity of various functional activities the last three or six months, e.g., domestic work as washing up after meals, gardening, household maintenance, working, and active participation in social activities. Each activity is scored on a Likert scale, ranging from 0 (never/ none) to 3 (weekly/ most days etc.). The scale has been psychometrically evaluated for elderly populations (Imam & Miller, 2012; McPhail et al., 2009).

### 2.3 Accelerometer based assessment of physical activity

Participants used an ankle-worn step activity measure (The Modus StepWatchTM3 Activity Monitor) for in median seven consecutive days, ranging from three to nine days. The protocol has been described elsewhere (Sanders et al., 2021). Briefly, the step activity measure recorded the stride count for every one-minute period for the whole test duration, and the recorded numbers were doubled to capture strides from both legs. Participants wore the StepWatch for minimum 600 minutes per day (mean = 859 minutes, SD = 67), omitting hours with zero counting for more than 90 consecutive minutes, with the possible exception of two minutes interruption. The 30 minutes before and after this interruption were required to be zero count. Mean total steps per day was analyzed together with metrics representing duration of low intensity (LPA) (> 0 steps/minute to < 100 steps/minute) and moderate-to-vigorous intensity (MVPA) (≥ 100 steps/minute) physical activity, based on a study linking oxygen consumption while walking with pedometer-based measures (Marshall et al., 2009). The results were averaged across all valid days of usage.

### 2.4 MRI acquisition

MRI was completed on a 3T MR 750 Discovery™ MRI scanner (GE healthcare, Milwaukee, USA) with a 32-channel head coil at the Oslo University Hospital, Oslo, Nrway.

We applied a Pseudo-Continuous ASL (PCASL) sequence with images obtained with an interleaved 3D spiral fast spin echo (FSE) readout module. Scan time was 4:55 minutes, with the following parameters: 512 sampling points on eight spirals, spatial resolution = 4 × 4 × 3 mm, default reconstructed spatial resolution = 2 × 2 × 3 mm, TR = 5025, TE = 11072 ms, labeling duration = 1450 ms, post labelling delay = 2025 ms, slice thickness = 3 mm, number of slices = 104, number of excitations = 3, and FOV = 256 mm. Care was taken when placing the lower edge of the 3D slab just below the cerebellum, which ensured that the distance to the labelling plane was approximately 9 cm below the anterior commissure to posterior commissure (AC-PC) line, in head to feet direction (Aslan et al., 2010).

Structural T1-weighted MRI were acquired using an inversion recovery-fast spoiled gradient echo (BRAVO) sequence with 188 sagittal slices. Scan time was 4:43 minutes, TE = 3.18 ms, TR = 8.16 ms, T1 = 450 ms, field of view (FOV) = 256 mm, acquisition matrix = 256 × 256, 1 mm^3^ isotropic voxels, flip angle (FA) = 12°.

### 2.5 MRI processing and analysis

ASL data were processed using FMRIB Software Library (FSL) (Jenkinson et al., 2012). Briefly, CBF maps were computed using Bayesian Inference for ASL MRI (BASIL) (Chappell et al., 2009), with the following parameters: tissue T1 = 1.2 s, arterial T1 = 1.6 s, labeling efficiency = 0.6, bolus arrival time = 1.45 s, bolus duration = 1.45 s, inversion time TI = 3.475 s, and blood-brain barrier coefficient 0.98 ml/g. A proton-density weighted calibration scan (M0) with equal readout as ASL and acquired in the same scan (the 40 last seconds), was used to voxel-wise estimate equilibrium blood magnetisation. Further, spatially smoothing of the CBF map was done utilizing an adaptive filter (Groves et al., 2009).

T1-weighted data was processed using FreeSurfer 5.3 (http://surfer.nmr.mgh.harvard.edu, Fischl, 2012), including automated segmentation and parcellation (Dale et al., 1999; Fischl et al., 2002). A visual quality control of all the reconstructed images was performed to exclude images with artefacts.

Using a previously described pipeline (Rokicki et al., 2021), individual CBF maps were co-registered to the reconstructed T1-weighted structural volumes using *bbregister* (Greve & Fischl, 2009). Individual Freesurfer-derived cortical and subcortical anatomical regions of interest (ROIs) were used to extract mean CBF from the accumbens area, amygdala, caudate, cerebellum cortex, hippocampus, pallidum, putamen and thalamus, in addition to the cerebral cortex, which was mapped across the surface using the Freesurfer-based cortical reconstructions. The ROIs were selected based on previous publications showing age-related associations with CBF (Chen et al., 2011; Stoquart-ElSankari et al., 2007; Zhang et al., 2017), as well as putative associations with everyday functioning (Sanchez et al., 2020) and daily physical activity (Zlatar et al., 2019). To decrease the number of comparisons, mean bilateral CBF for each anatomical region was included in the further analysis.

### 2.6 Activity decomposition

Prior studies have suggested that FAI captures three underlying dimensions: domestic, work/ leisure and outdoor activities (Bond et al., 1992; Cockburn et al., 1990). Accordingly, we performed an exploratory factor analysis based on polychoric correlation to reduce the amount of data for further analysis. Due to low variability, the items “Walking outside for > 15 minutes” and “Driving car/ going on bus” were excluded from further analysis. The items “Gainful work” and “Actively pursuing hobby” were dichotomized to “yes” or “no” and “never to less than weekly” or “at least weekly”, respectively, and excluded from the factor analysis. The items “Reading books”, “Gardening” and “Household maintenance” were also excluded from the factor analysis due to low correlation with the rest of the items (“Gardening”), and a factor loading of < 0.5 (“Reading books” and “Household maintenance”). The correlation structure among the remaining items confirmed commonalities (Supplementary Figure 1). Principal axis factor analysis was conducted with orthogonal rotation (oblimin). Two factors were considered, as suggested by parallel analysis. Factor 1, named “Domestic work” comprised five items explaining 44.3 % of the variance with factor loadings from 0.61 to 0.85, and factor 2, named “Social activities” comprised two items explaining 10.9 % of the variance with factor loadings from 0.52 to 0.68 (see Supplementary Table 1 for additional information).

### 2.7 Statistical analysis

Statistical analyses were performed using R, version 3.6.2 (R Core Team, 2019). For descriptive purposes, steps per day, including number of steps taken at different intensities (low and moderate-to-vigorous), and body mass index (BMI) are reported. Correlations between the activity measures, the CBF measures, and age were analyzed using Kendall τ or Pearson’s r, as appropriate.

We addressed the main hypotheses of associations between baseline and follow-up CBF and various activity measures using Bayesian multilevel linear models using Stan (Stan Development Team, 2020) and the *brms* packages in R (Bürkner, 2017). The association between baseline CBF and activity level was assessed by applying the CBF measures as dependent variables with each measure of activity level independently used together with age and sex as independent variables (CBF ∼ activity level + age + sex). The latter were included due to previous studies suggesting an association with CBF (Alisch et al., 2021). Since several models revealed moderate to strong evidence of an association between sex and CBF, explorative analyses were completed including an interaction term between sex and activity measures (CBF ∼ activity level × sex + age). Analyzing the association between CBF and FAI data, one case was considered an outlier with possible relatively large influence on the regression line. Regression diagnostics for influential cases suggested a Cook’s distance greater than 3 times the mean, and as a consequence, the case was removed from further baseline analysis concerning FAI. Analyzing the interaction effect of sex on the association between baseline MVPA and CBF suggested evidence of an interaction effect mainly driven by six females, all considered outliers with MVPA-level of more than two standard deviations above mean (approximately one hour MVPA daily). No males had this high level of average daily minutes of MVPA. Regression diagnostics for influential cases suggested a Cook’s distance greater than 3 times the mean, on multiple of the baseline analysis including MVPA, for five of the six cases. As a consequence, and to promote generalizability, the six cases were removed from baseline analysis considering MVPA (Supplementary Figure 2).

Using Bayesian mixed effects modelling, we assessed the hypothesis that higher activity score at baseline was associated with less decline in CBF between the two MRI assessments. Data from the two timepoints of each CBF region was used as dependent variables in the models, and separate models were run for each activity measure. Each model was adjusted for baseline age and sex. Random intercepts for each subject were entered into the models as random effects (CBF ∼ activity measure × timepoint + age + sex + (1|subject)). Prior probability distributions were set to zero with a standard deviation of 0.5 for all parameters. All continuous variables in the models were scaled to have a mean of zero, and a standard deviation of one, prior to running the analysis. Due to low variability in the 86 participants completing the follow-up scan, the item “hobby” was not included in the longitudinal analysis (16 participants scoring 0 = “never to less than weekly”, and 70 participants scoring 1 =“at least weekly”). Results were presented with mean estimated coefficient, 95% credible interval using highest density interval (HDI) method, and Bayes Factor (BF). BF was calculated as evidence for the null or alternative hypothesis with the use of Savage-Dickey method (Wagenmakers et al., 2010). For a full overview of the evidence categories, see Supplementary Figure 3. Briefly, BF > 1 indicates evidence of the null hypothesis, BF < 1 indicates evidence of the alternative hypothesis, and BF = 1 indicates no evidence in any directions.

## 3. Results

### 3.1 Sample characteristics

Table 1 and Figure 1 summarize sample characteristics, Figure 2 shows distributions of the baseline activity measures for females and males, and Figure 3 shows the pairwise correlation matrix among the activity and CBF measures, respectively, as well as age and BMI. At baseline, 48 % were considered active according to physical activity definitions of 150 minutes of MVPA per week (World Health Organization, 2020), here pragmatically operationalized as >20 minutes per day. The participants completed on average 12,084 (SD = 3643) steps per day, of which an average of 78 % were defined as low-intensity and 22 % moderate-to-vigorous intensity steps. Participants were on average active for 384 minutes (SD = 79) per day, including 94 % in the low-intensity range and 6 % in the moderate-to-high intensity range. Domestic work, social activities, maintenance, and different leisure activities were reported as regular activities. 71 % of the participants were not working, and 21 % were pursuing leisure activities and hobbies less than weekly. In general, the associations between the different activities, including BMI, were low, with exception of gardening and maintenance (τ = 0.44, *p* < .001), and domestic work and social activity (τ = 0.35, *p* < .001).

**Table 1.**
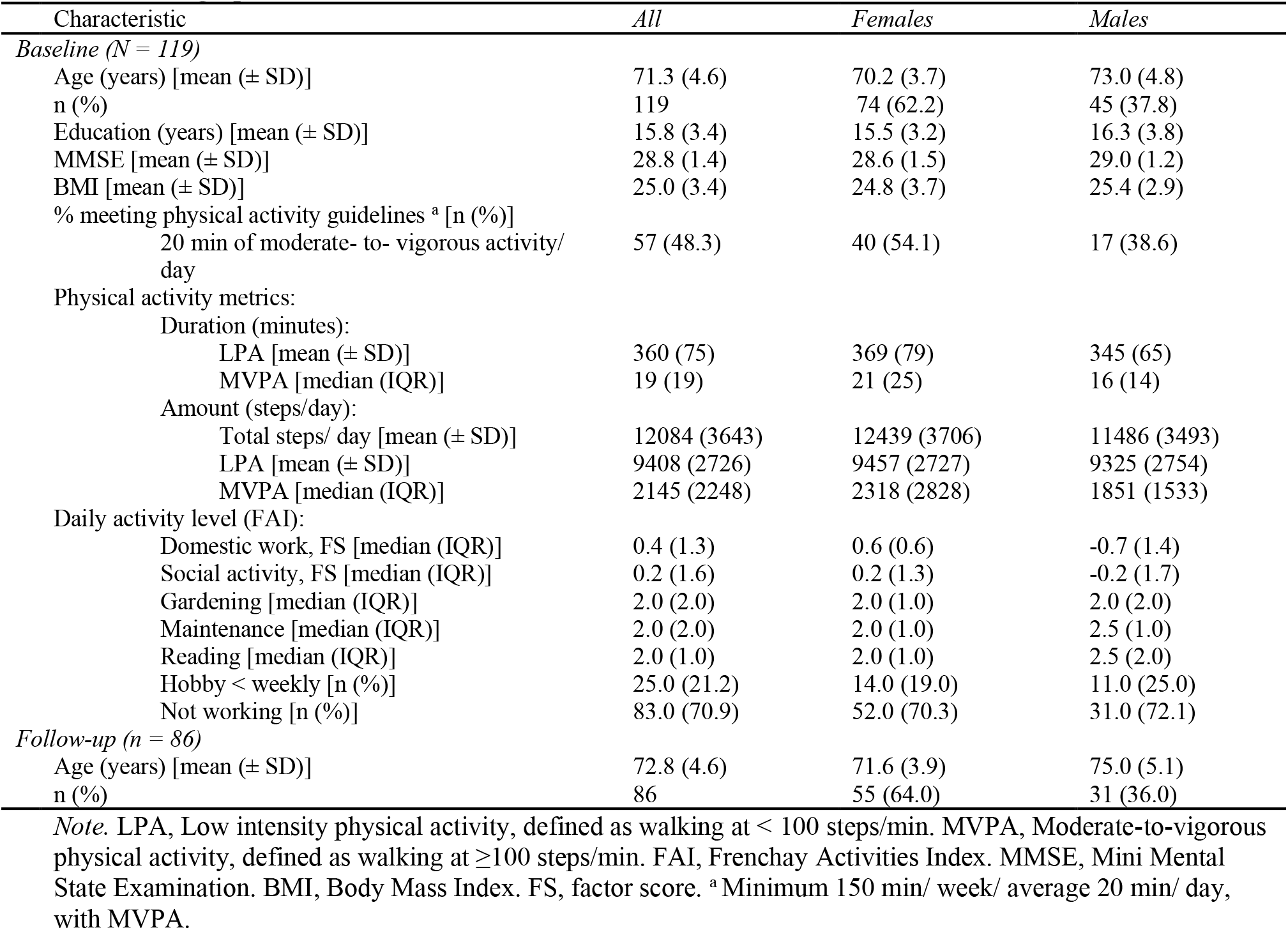
Demographics and clinical characteristics.

| Characteristic | All | Females | Males |
| --- | --- | --- | --- |
| *Baseline (N = 119)* |  |  |  |
| Age (years) [mean (± SD)] | 71.3 (4.6) | 70.2 (3.7) | 73.0 (4.8) |
| n (%) | 119 | 74 (62.2) | 45 (37.8) |
| Education (years) [mean (± SD)] | 15.8 (3.4) | 15.5 (3.2) | 16.3 (3.8) |
| MMSE [mean (± SD)] | 28.8 (1.4) | 28.6 (1.5) | 29.0 (1.2) |
| BMI [mean (± SD)] | 25.0 (3.4) | 24.8 (3.7) | 25.4 (2.9) |
| % meeting physical activity guidelines <sup>a</sup> [n (%)] |  |  |  |
| 20 min of moderate- to- vigorous activity/ day | 57 (48.3) | 40 (54.1) | 17 (38.6) |
| Physical activity metrics: |  |  |  |
| Duration (minutes): |  |  |  |
| LPA [mean (± SD)] | 360 (75) | 369 (79) | 345 (65) |
| MVPA [median (IQR)] | 19 (19) | 21 (25) | 16 (14) |
| Amount (steps/day): |  |  |  |
| Total steps/ day [mean (± SD)] | 12084 (3643) | 12439 (3706) | 11486 (3493) |
| LPA [mean (± SD)] | 9408 (2726) | 9457 (2727) | 9325 (2754) |
| MVPA [median (IQR)] | 2145 (2248) | 2318 (2828) | 1851 (1533) |
| Daily activity level (FAI): |  |  |  |
| Domestic work, FS [median (IQR)] | 0.4 (1.3) | 0.6 (0.6) | -0.7 (1.4) |
| Social activity, FS [median (IQR)] | 0.2 (1.6) | 0.2 (1.3) | -0.2 (1.7) |
| Gardening [median (IQR)] | 2.0 (2.0) | 2.0 (1.0) | 2.0 (2.0) |
| Maintenance [median (IQR)] | 2.0 (2.0) | 2.0 (1.0) | 2.5 (1.0) |
| Reading [median (IQR)] | 2.0 (1.0) | 2.0 (1.0) | 2.5 (2.0) |
| Hobby < weekly [n (%)] | 25.0 (21.2) | 14.0 (19.0) | 11.0 (25.0) |
| Not working [n (%)] | 83.0 (70.9) | 52.0 (70.3) | 31.0 (72.1) |
| *Follow-up (n = 86)* |  |  |  |
| Age (years) [mean (± SD)] | 72.8 (4.6) | 71.6 (3.9) | 75.0 (5.1) |
| n (%) | 86 | 55 (64.0) | 31 (36.0) |

*Note.* LPA, Low intensity physical activity, defined as walking at < 100 steps/min. MVPA, Moderate-to-vigorous physical activity, defined as walking at ≥ 100 steps/min. FAI, Frenchay Activities Index. MMSE, Mini Mental State Examination. BMI, Body Mass Index. FS, factor score. <sup>a</sup> Minimum 150 min/ week/ average 20 min/ day, with MVPA.

**Figure 1.**
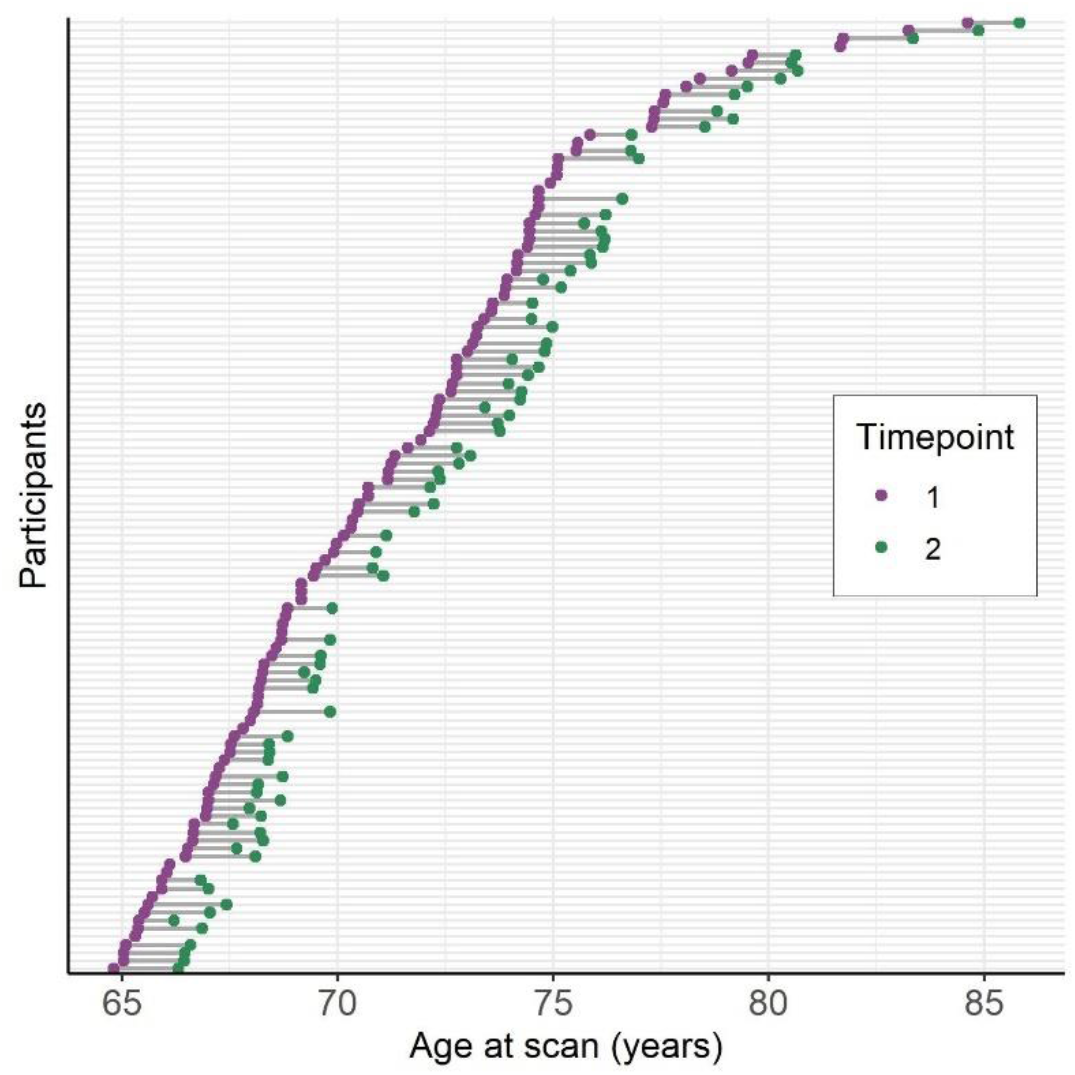
Age distribution for all participants and all available data where purple points denote a scan acquisition included in the analysis at baseline (*N* = 119) and green points denote age at follow-up (*n* = 86). Two scans of the same participant are denoted by the line connecting the dots. The participants are sorted by age at baseline and the y-axis represents the individual participants.

**Figure 2.**
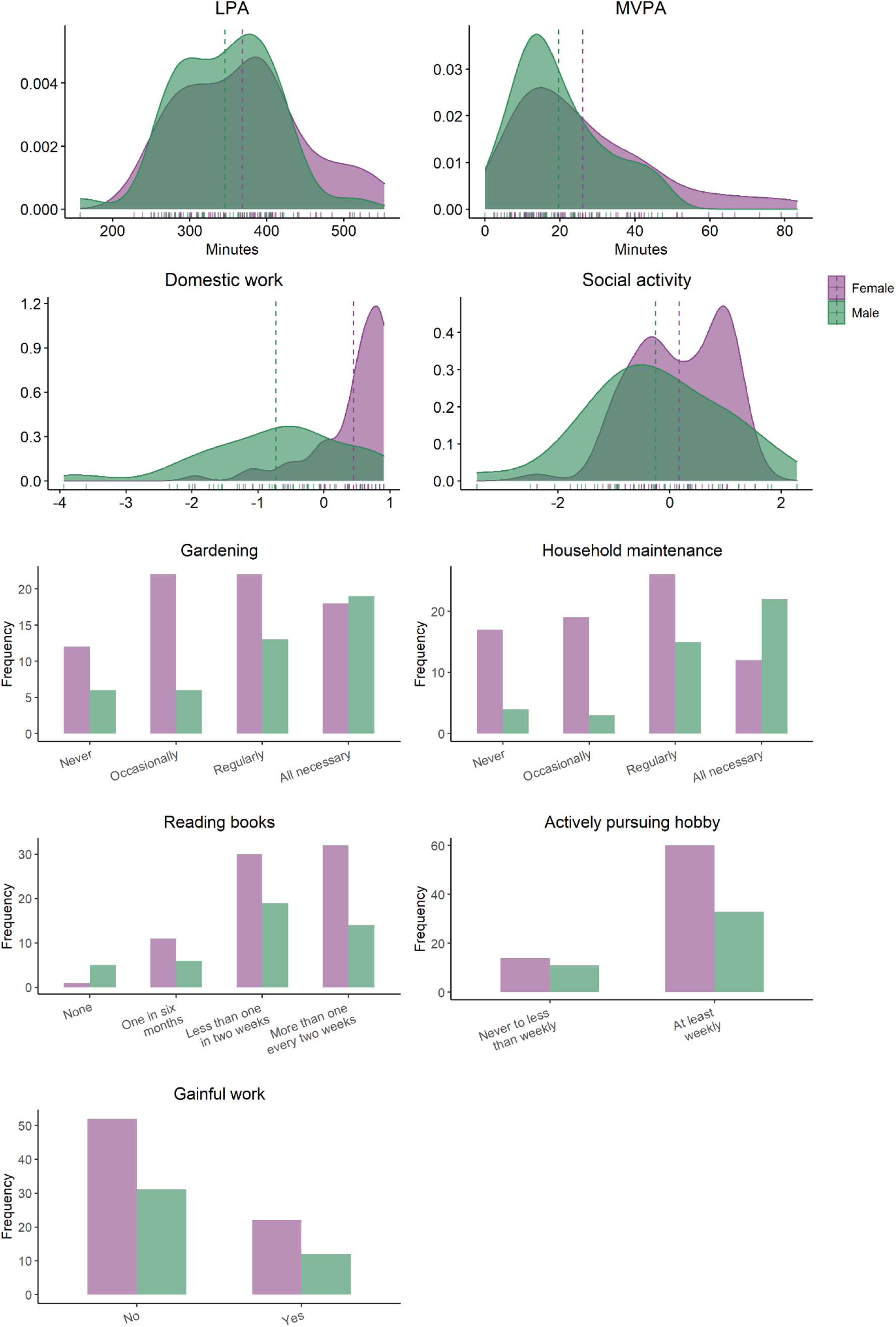
Distribution of baseline activity measures (*N* = 119) split by sex. Density plot with vertical lines depict mean values for each sex. LPA: Average daily minutes with low intensity physical activity (< 100 steps per minute), MVPA: Average daily minutes with moderate to vigorous intensity physical activity (≥ 100 steps/minute).

**Figure 3.**
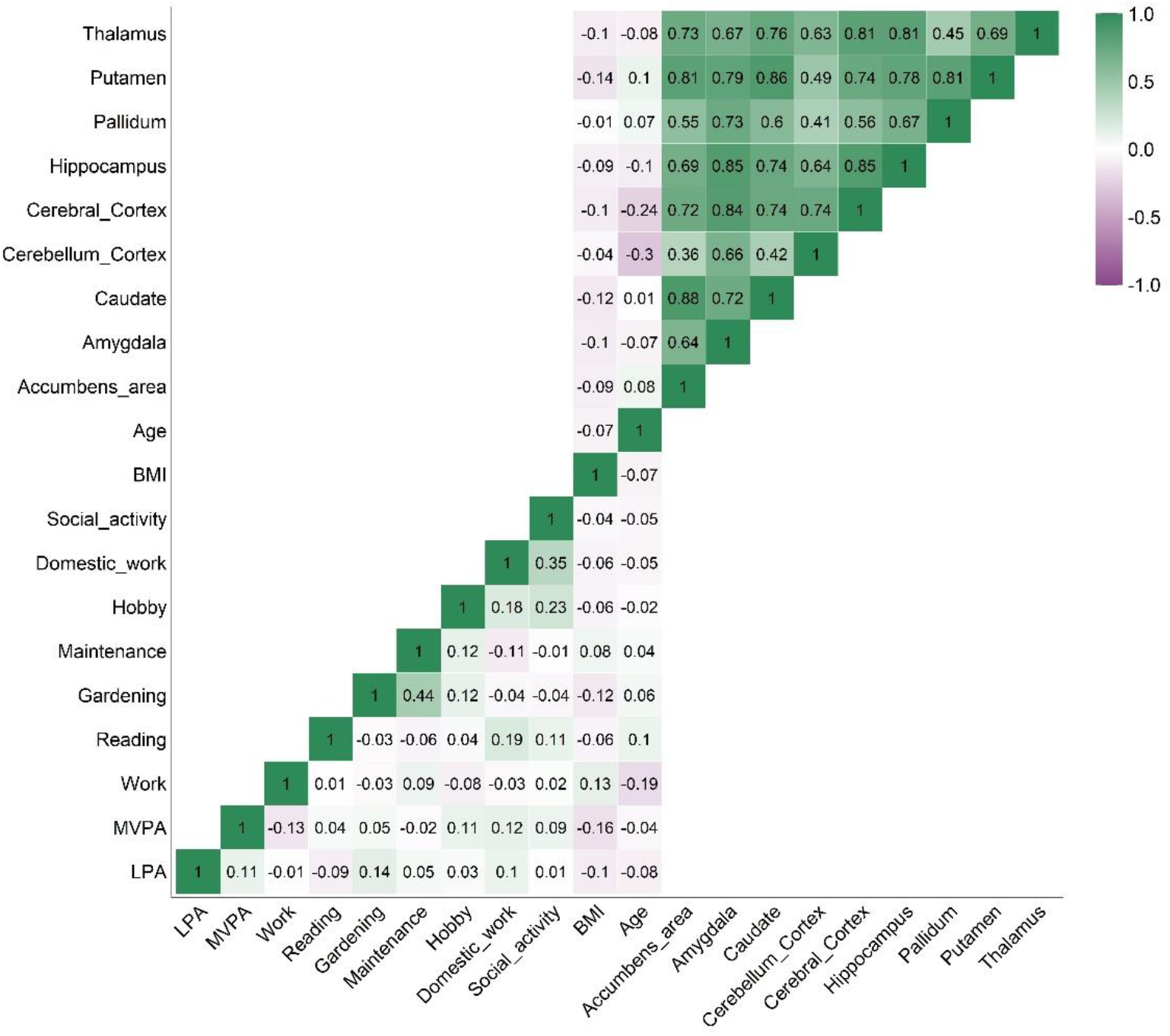
Correlation matrix of baseline physical activity measures (Kendall τ), regional cerebral blood flow (Pearson’s r), BMI and age.

There were several strong positive associations among the baseline regional CBF measures, and moderate negative associations between age and cortical CBF (r = -0.24, p = .007) and cerebellar cortical CBF (r = -0.30, p < .001), while the remaining associations, including between CBF and BMI, were low.

Table 2 summarizes descriptive data on CBF, Figure 4 presents plots of mean regional CBF for the two timepoints and association between estimated yearly change in CBF and age, and Supplementary Figure 4 shows the pairwise correlation matrix on estimated yearly change in CBF. For the 86 participants with both baseline and longitudinal CBF, mean (min-max) intraclass correlation coefficient (ICC) for all the CBF measures was 0.49 (0.35 - 0.59), indicating moderate reliability between the two timepoints. Median estimated yearly change in CBF ranged between -0.2 (SD = 5.4) ml/100 g/min for putamen to -1.4 (SD = 7.0) ml/100 g/min for cerebellum cortex. Estimated yearly change for CBF accumbens was negatively correlated with age (τ = -0.17, *p* = .019), indicating larger reduction in CBF with higher age. For the remaining ROIs, the associations between annual change in CBF and age were low. Correlation between estimated yearly change in CBF range from r = 0.25 (CBF pallidum and thalamus) to r = 0.85 (CBF caudate and accumbens).

**Table 2.** Descriptive data on CBF

| | Correlation between age and baseline CBF ($r$ [p]) | EYC in CBF (median [IQR]) | Correlation between age and EYC CBF ($\tau$ [p]) | Main effect of time on CBF ($\beta$/ HDI/ BF)<sup>†</sup> | ICC (CI/ F/ p) |
| --- | --- | --- | --- | --- | --- |
| Cerebral cortex | -0.24 (.007)* | -0.6 (6.7) | -0.12 (.102) | -0.12/ -0.32, 0.08/ 5.0 | 0.59 (0.43, 0.71/ 3.84/ < .001*) |
| Cerebellum cortex | -0.30 (.001)* | -1.4 (7.0) | -0.06 (.438) | -0.14/ -0.36, 0.06/ 3.82 | 0.53 (0.36, 0.67/ 3.27/ < .001*) |
| Accumbens area | 0.08 (.374) | -1.0 (7.0) | -0.17 (.019)* | -0.03/ -0.28, 0.21/ 7.73 | 0.35 (0.15, 0.52/ 2.08/ < .001*) |
| Amygdala | -0.07 (.437) | -0.5 (4.6) | -0.04 (.591) | -0.08/ -0.3, 0.13/ 6.94 | 0.51 (0.33, 0.65/ 3.07/ < .001*) |
| Caudate | 0.01 (.955) | -0.2 (5.5) | -0.10 (.184) | 0.0/ -0.23, 0.23/ 8.72 | 0.45 (0.26, 0.60/ 2.64, < .001*) |
| Hippocampus | -0.10 (.304) | -0.4 (5.8) | -0.13 (.085) | -0.07/ -0.29, 0.15/ 7.42 | 0.47 (0.29, 0.62/ 2.79/ < .001*) |
| Pallidum | 0.07 (.427) | -0.2 (5.3) | -0.05 (.536) | -0.03/ -0.23, 0.19/ 9.26 | 0.52 (0.35, 0.66/ 3.17/ < .001*) |
| Putamen | 0.10 (.297) | -0.2 (5.4) | -0.14 (.059) | 0.04/ -0.16, 0.25/ 8.60 | 0.54 (0.37, 0.67/ 3.34/ < .001*) |
| Thalamus | -0.08 (.407) | -0.6 (6.5) | -0.13 (.066) | -0.17/ -0.4, 0.08/ 2.99 | 0.41 (0.12, 0.57/ 2.36, < .001*) |

*Note.* IQR, Inter quartile range; $\beta$ , estimate; HDI, highest density interval; BF, Bayes Factor; EYC, estimated yearly change; ICC, Intraclass correlation coefficient; CI, Confidence interval; F, F-test, \* $p < .05$ <sup>†</sup>Bayes mixed effect model: CBF ~ Age + Sex + (1|Subject)

**Figure 4.**
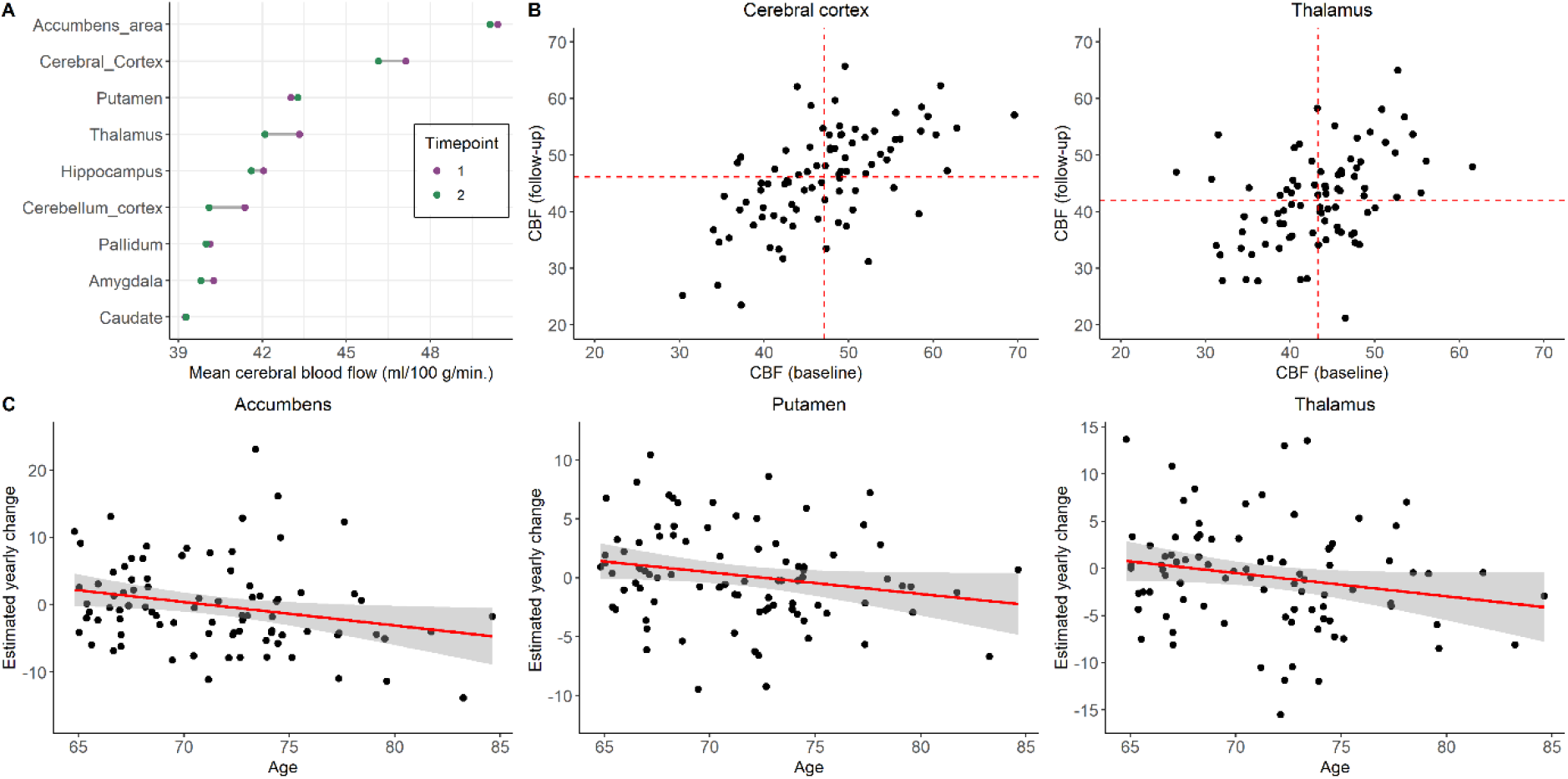
(A) Mean cerebral blood flow (CBF) where purple points denote mean CBF at baseline *(N* = 119) and green points denote mean CBF at follow-up (*n* = 86). Two scans of the same structure are denoted by connecting line. (B) Scatter plots of CBF from the longitudinal sample (*n* = 86) at baseline and follow-up for cerebral cortex and thalamus, (C) and examples of association with fit line between estimated yearly change (EYC) in CBF and age at baseline (*n* = 86).

### 3.2 Associations between CBF and frequency of activities

Figure 5 summarizes mean parameter estimates and credible intervals for the associations between baseline CBF and activity level. Supplementary Figure 5 presents evidence ratios for the same associations, and Supplementary Table 2 presents summary statistics. The analysis revealed moderate evidence for a positive association between accumbens CBF and minutes of moderate-to-high intensity physical activity (> 100 steps/min) (β = 0.31, HDI = 0.08, 0.53, BF = 0.13), a positive association with pallidum CBF and minutes of low intensity physical activity (β = 0.24, HDI = 0.05, 0.41, BF = 0.22), a positive association for thalamic CBF and reading (β = 0.27, HDI = 0.07, 0.48, BF = 0.17), and a negative association between thalamic CBF with maintenance (β = -0.23, HDI = -0.04, -0.06, BF = 0.20). In addition, the analysis suggested anecdotal evidence for a positive association between accumbens CBF and low intensity physical activity level (< 100 steps/min) (β = 0.18, HDI = 0.0, 0.36, BF = 0.85), a negative association between accumbens CBF and work (β = -0.35, HDI = -0.71, -0.03, BF = 0.47), and a positive association between caudate CBF and minutes of moderate-to-high intensity physical activity (β = 0.22, HDI = -0.03, 0.45, BF = 0.80). Anecdotal evidence also supported a positive association between cortical CBF and reading (β = 0.19, HDI = 0.0, 0.39, BF = 0.78), a positive association between cortical CBF and participation in social activity (β = 0.17, HDI = 0.01, 0.34, BF = 0.83), and for a positive association between pallidum CBF and hobby (β = 0.30, HDI = -0.01, 0.71, BF = 0.87). For putamen CBF, anecdotal evidence of a positive associations with low intensity physical activity level (β = 0.20, HDI = 0.02, 0.38, BF = 0.49) and negative association with maintenance (β = -0.18, HDI = -0.36, -0.01, BF = 0.69) were found. Figure 6 shows scatter plots of the models with evidence suggesting an association between CBF and activity level.

**Figure 5.**
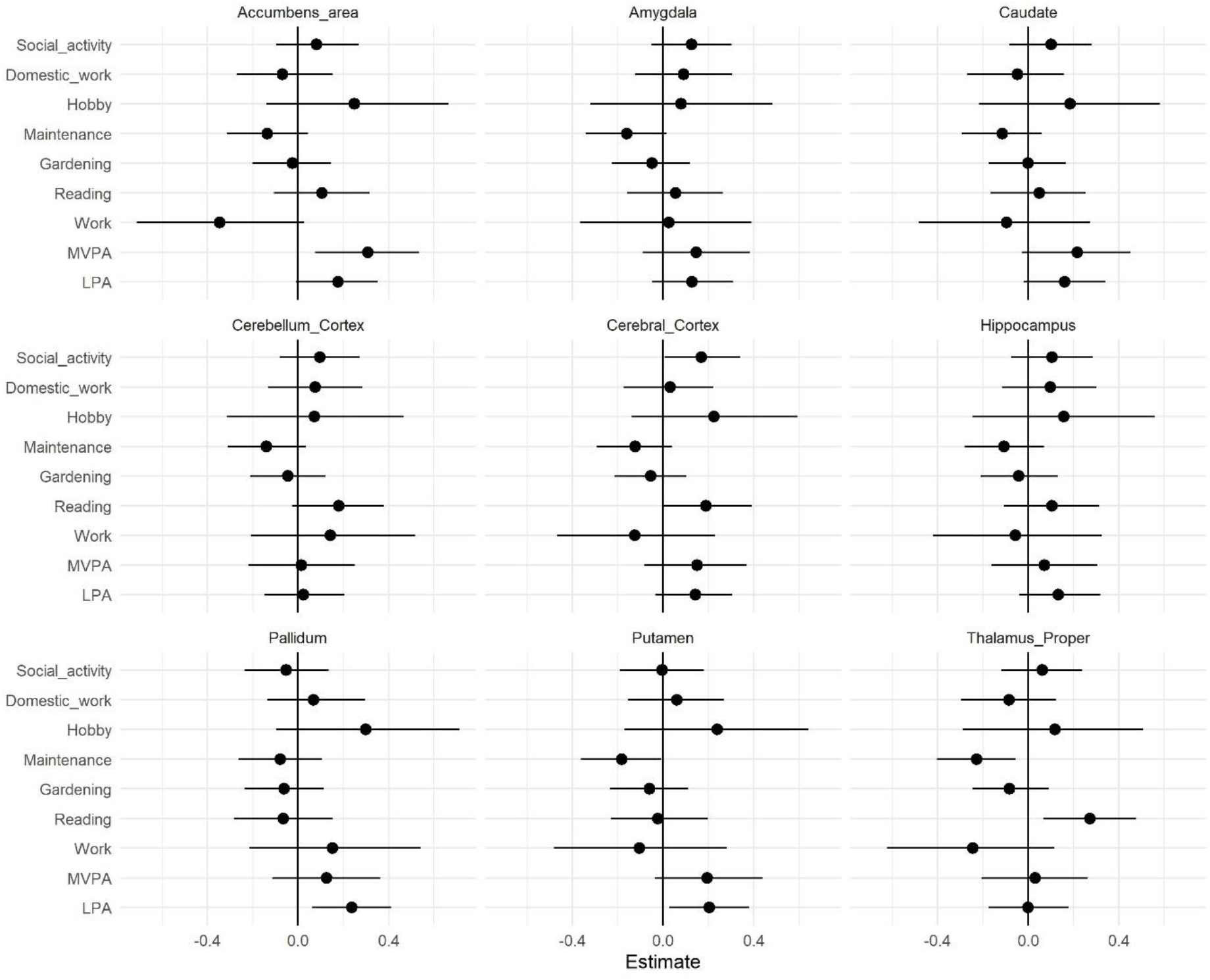
Parameter estimates for the association between baseline CBF and measures of activity level with 95% credible interval.

**Figure 6.**
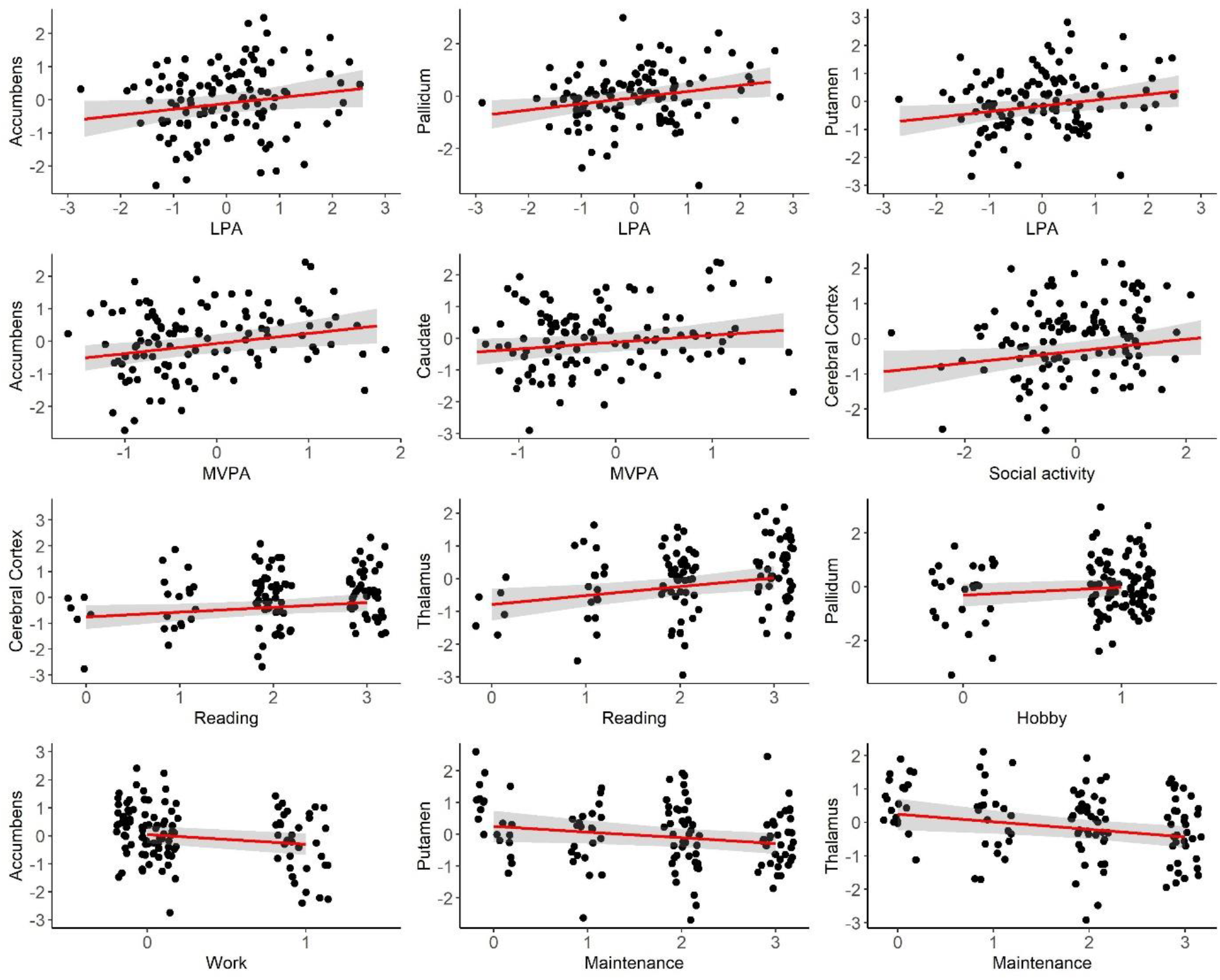
Associations between regional CBF and activity level. Points represent observed data. Regression lines represent estimated slopes and grey band represents 95 % credible interval (uncertainty). The numeric data has been scaled before analysis. LPA: Average daily minutes with low intensity physical activity (< 100 steps per minute), MVPA: Average daily minutes with moderate to vigorous physical activity (≥ 100 steps/minute).

Moderate to strong evidence of a main effect of sex on CBF were found across several models, with higher CBF among women compared to men (Supplementary Table 3). Secondary analyses revealed no evidence of an interaction effect between sex and activity measures on regional CBF (Supplementary Figures 6 and 7, Supplementary Tables 4 and 5). When including an interaction between sex and activity, anecdotal evidence of a positive association between cortical CBF and frequency of hobbies was found (β = 0.31, HDI = -0.11, 0.72, BF = 0.80).

### 3.3 Interaction effect between time and frequency of activities on CBF

Figure 7 summarizes parameter estimates and corresponding credible intervals for the associations between longitudinal changes in CBF and activity level. The analysis revealed no evidence of an association between baseline level of activity and change in regional CBF. For evidence ratios and summary statistics, please refer to Supplementary Figure 8 and Table 6.

**Figure 7.**
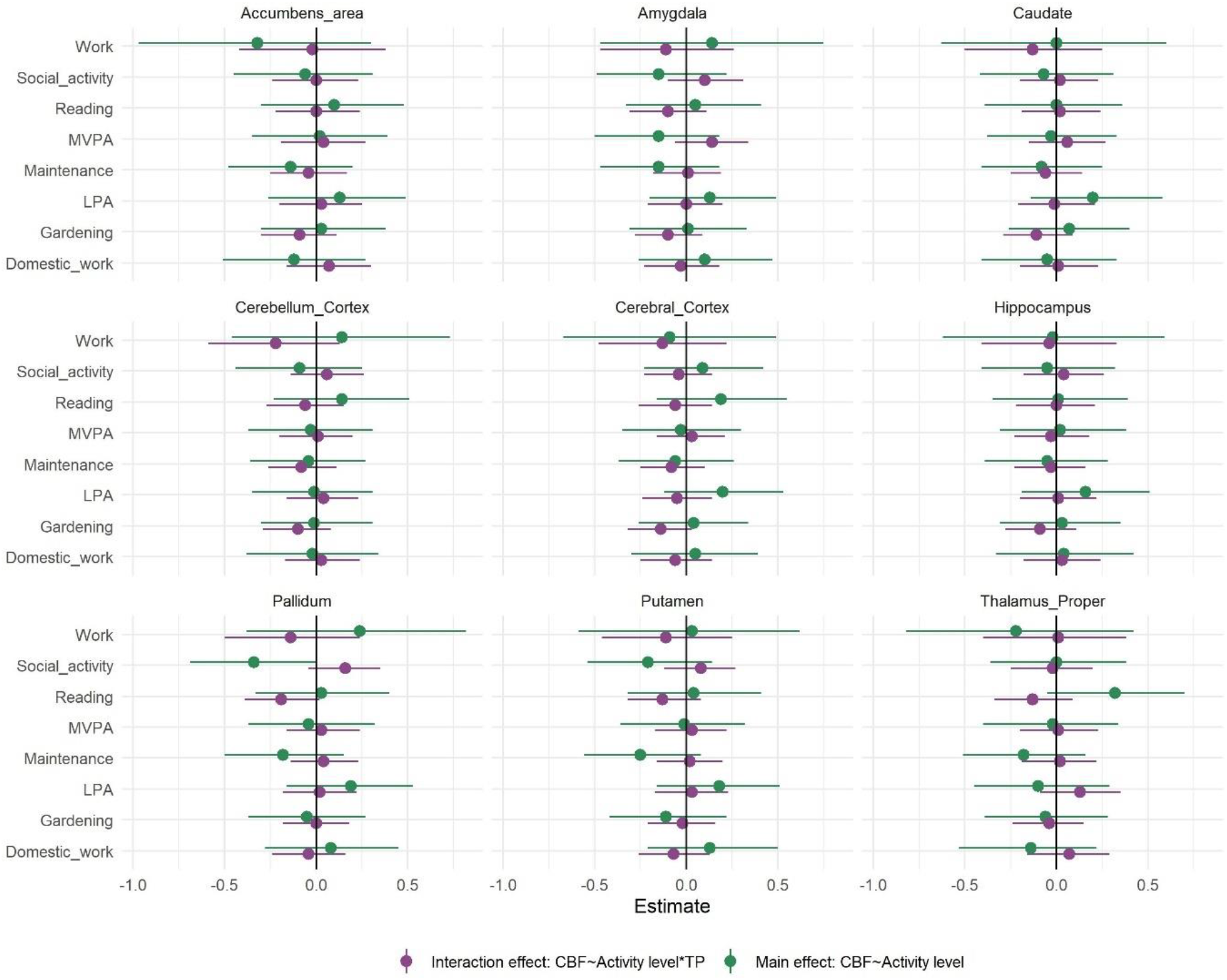
Parameter estimates reflecting the interaction effect between timepoint and frequency of activities on CBF (purple) and the association between regional CBF and activity level (green) with 95 % credible interval. Model: CBF∼ activity measure × timepoint + age + sex + (1|subject).

When including an interaction term between timepoint and activity level, anecdotal evidence of a positive association across time points between thalamic CBF and reading was found (β = 0.32, HDI = -0.05, 0.70, BF = 0.64), in addition to negative associations between accumbens CBF and work (β = -0.32, HDI = -0.97, 0.30, BF = 0.95), pallidum CBF and social participation (β = -0.34, HDI = -0.69, 0.0, BF = 0.43) and putamen CBF and maintenance (β = -0.25, HDI = -0.56, 0.08, BF = 0.93) (Supplementary Table 7).

## 4. Discussion

Intact cerebrovascular function is an important constituent of healthy cognitive and brain aging. Identifying correlates and putative modifiers of age-related changes in cerebral blood flow before the emergence of incipient clinical decline may inform public health advice and clinical practice. The present study is one of the first studies of the association between level of everyday activities and cortical and subcortical CBF in older adults, including both cross-sectional and longitudinal assessments, objective measures and definitions of low, moderate-to-vigorous intensity physical activity, and frequency of specific everyday activities. Bayesian analyses indicated anecdotal to moderate associations between both cortical and subcortical CBF and various activities, including between higher baseline CBF and higher level of low, moderate-to-intensive physical activity and various leisure activities. In contrast, the analyses revealed no evidence of a robust association between baseline activity level and CBF changes between two assessments across the follow-up period.

Prior studies examining the importance of cortical and subcortical CBF for maintaining cognitive functions in ageing have showed higher CBF in multiple regions in younger compared to older healthy individuals (Chen et al., 2011; Stoquart-ElSankari et al., 2007; Zhang et al., 2017). Lower CBF is considered to be a risk factor for development of age-related neurocognitive disorders including dementia (de la Torre, 2012), suggesting that protecting the brain against age-related changes in CBF is beneficial for future cognitive functioning and outcome. In the present study we show that more time spent on low intensity physical activity is associated with higher CBF in multiple subcortical regions of the brain, including the accumbens, putamen and pallidum. These basal ganglia regions are vulnerable to the adverse effects of small vessel disease (Pantoni, 2010), and are implicated in learning, motivation, coordination and regulation of motor movements (Gazzaniga et al., 2013). The results complement prior cross-sectional associations between objectively measured low intensive physical activity and CBF in frontal regions among 52 cognitive healthy older adults (Zlatar et al., 2019). While causal interpretations should be made with caution, these cross-sectional findings collectively support the assumption that higher level of low intensive physical activity, in addition to exercise and overall fitness, may counteract age-related neurovascular changes. Moreover, previous intervention studies have also reported associations between exercise and regional CBF (Chapman et al., 2013; Kleinloog et al., 2019), in line with our observational study that also found evidence for an association between subcortical CBF and minutes of moderate-to-vigorous physical activity. The positive associations between both low and moderate-to-intensive physical activity and subcortical CBF corroborate previous converging evidence suggesting cognitive and other health-related benefits of maintaining physical activity throughout adulthood, including higher quality of life, physical and cognitive function, fewer depressive symptoms (Buchman et al., 2019; Stubbs et al., 2017; Varma et al., 2014), and also increased functional connectivity in higher-level cognitive brain networks (Voss et al., 2010). Importantly, the latter association was found independent of post-mortem brain pathology and regardless of intensity of the daily activity.

The positive association between time spent on physical activity and subcortical CBF may be explained by different mechanisms, but studies are scarce, and mostly comprise animal studies and exercise (Davenport et al., 2012; Zimmerman et al., 2014). Daily low intensive physical activity can be considered among activities that accumulate daily energy expenditure and maintain a certain muscular strength. Furthermore, physical activity may help prevent the development of frailty (Oliveira et al., 2020), which is an increasingly recognized risk factor for adverse brain health outcomes (Gallucci et al., 2022). Despite the low intensity, some of the same mechanisms explaining the positive effect of exercise on CBF may apply. It has been shown that increased blood flow in response to physical exercise increases vascular shear stress, which in turn might lead to an upregulation of endothelial nitric oxide (NO) synthase expression, important for NO dependent vasodilation, and further an increase in basal CBF (Endres et al., 2003). Age-related reduction in CBF may also directly influence energy and metabolic supply by hindering important nutrients to reach areas with metabolic demands, and contribute to neurodegenerative changes (Davenport et al., 2012). Further, an upregulation of CBF may have a neuroprotective effect. Multiple neurotropic factors have been suggested upregulated with exercise, contributing to both angiogenesis and neurogenesis, and includes vascular endothelial growth factor (VEGF), insulin-like growth factor 1 (IGF-1) and brain-derived neurotrophic factor (BDNF), with potential neuroprotective effect and contributors to neurogenesis (Bjørnebekk et al., 2005; Cotman et al., 2007). Animal studies have provided knowledge on the effect of physical exercise on both neurogenesis (van Praag et al., 1999; Voss et al., 2013) and angiogenesis (Davenport et al., 2012; Swain et al., 2003). However, the molecular mechanisms are still uncertain. A recent animal study revealed an upregulation of 68 proteins in blood plasma levels in response to a running intervention compared to sedentary controls, and suggested that the effect of increasing neurogenesis in the hippocampus in relation to exercise is particularly supported by the release of antioxidant selenium transport protein (Leiter et al., 2022). Increase in oxidative stress and endothelial dysfunction in older adults has been suggested to reduce cerebrovascular reactivity to hypercapnia, with increase in antioxidants as an important contributor protecting the vasculature from damage (Davenport et al., 2012).

Importantly, animal studies demonstrating neuroprotective effects of a running intervention found similar results when the mice were exposed to an enriched living standard, including social participation, opportunities for learning, larger environments and in general more physical activity (van Praag et al., 1999), indicating overall positive effects of an enriched physical and social environment on the brain. The current analysis revealed a positive association between subcortical and cortical CBF and frequency of more cognitive demanding leisure activities, reading, engaging in hobbies and social activities. A previous longitudinal study demonstrated that while the use of computer for > 1 hour daily was associated with better cognitive function with advancing age, watching television was not (Kesse-Guyot et al., 2012). This is consistent with our findings, indicating that some everyday activities may indicate or be beneficial for cognitive function and brain health in older adults, but not necessarily all types of activity. Our cross-sectional results complement a previous longitudinal study including 1463 participants aged > 65 years showing decreased risk of cognitive decline with higher participation in leisure activities across a 2.4 years interval.

Higher frequency of mental activities, including reading and various hobby activities, were associated with global cognition, language and executive function, and social activities were associated with global cognition (Wang et al., 2013). Our findings also add to a previous longitudinal study of 942 participants showing reduced risk for developing dementia with higher level of commitment to hobbies (Hughes et al., 2010). In addition, social isolation has been associated with increased mortality, with risk factors including lower socioeconomic status and less healthy lifestyle (Elovainio et al., 2017).

With more cognitive demanding activities, such as reading, cognitive demanding hobbies, or social activities, there would potentially be higher demand on both neural processing, synaptic organization, and efficient and plastic neurological processes (Park & Bischof, 2013). As reviewed above, it is possible that neurogenesis increases the metabolic requirements and potentially promotes angiogenesis (Davenport et al., 2012; Swain et al., 2003), explaining the current cross-sectional associations between CBF and engaging in more cognitively demanding activities. Importantly, more social activities have been associated with more cognitive challenges and more physical movement (Wang et al., 2013), possibly explaining the current results. Further, the current questionnaire item engagement in hobbies could potentially include a variety of activities, from spending time outdoors to more sedentary activities such as knitting. To which degree the associations can be explained by physical versus cognitive mechanisms is unknown and needs further exploration. Regardless, the current results indicating that regular participation in easily accessible activities is associated with higher CBF are encouraging.

The results also indicated an association between higher frequency of maintenance and lower CBF in multiple subcortical regions. While the participation in home maintenance was higher for male than female participants, the analysis revealed no interaction effect with sex, suggesting similar associations among females and males. It is possible that participants spending more time on work or maintenance work did less of other health promoting behavior. The results highlight the complexity of exploring the health benefits of everyday activity participation and calls for further research.

The current analyses did not support an association between baseline activity level and longitudinal change in cortical or subcortical CBF. This complements the results from a 5-year randomized controlled trial, showing few differences in various brain imaging measures and intensity of physical activity (Pani et al., 2022; Pani et al., 2021). The participants in the present study can be considered highly active, with an average of > 12,000 steps per day as measured using the accelerometer, and with an average of 48 % fulfilling the current criteria for adults over the age of 65 of more than 150 minutes of moderate-to-vigorous activity per week (World Health Organization, 2020). None of the included participants were considered sedentary as defined by walking < 5,000 steps/ day (Tudor-Locke et al., 2013). In addition to including participants with lower activity level, the relative short follow-up period might have concealed possible associations, and a longer follow-up period can also be suggested for future studies.

Several limitations should be considered when interpreting the results of the current study. The generalizability of the results is influenced by the sample consisting of many active, healthy, well-educated, older adults. Future studies recruiting a greater span of individuals, including people with cerebrovascular diseases as well as associated comorbidities such as type-2 diabetes, obesity and ischemic heart disease, might contribute to increasing the generalizability of the findings. While the use of a reliable sensor to measure minutes of daily physical activity (Foster et al., 2005) is eminent at capturing steps, the accelerometer does not capture movements of the upper extremity, known to be an important part in physical activity, or discriminate between individual activity types. Further, potential seasonal variation in physical activity (Aspvik et al., 2018) was not controlled for in the current analysis. By including the FAI, encompassing an array of activities such as domestic work, gardening and hobbies, we ensured a wider examination of the concept of everyday activity level. While it is difficult to assess free-living activity level using a randomized controlled design (Buchman et al., 2019), the current observational study design does not allow for causal inference. Also, with only baseline measure of activity level, the positive associations between CBF and various everyday activities may result both from present activity level and from the accumulated effects across several years or even a lifespan. Intake of alcohol, caffeine and certain food types may give an acute effect on CBF. However, the long-term effects are less clear (Clement et al., 2018; Joris et al., 2018). Future studies may be able to integrate detailed measures of risk and health-related behaviors such as smoking, alcohol and drug use, as well as social measures such as loneliness, with activity measures.

In conclusion, with the ultimate aim of identifying potential targets for interventions, the primary objective of the present study was to test for associations between frequency of diverse everyday activities and cross-sectional and longitudinal measures of regional CBF among healthy community-dwelling adults aged 65-89 years. The findings support a positive association between CBF and regular participation in various leisure activity, including low and moderate-to-vigorous intensity physical activity, social participation, hobbies, and reading. Although baseline level physical activity was not associated with further CBF changes across the one-to-two years interval between assessments, the current work provides evidence of a general positive association between a physically active and enriched lifestyle and cerebral vascular functions in elderly participants, which may point to candidate target for interventions in future clinical trials.

## Supporting information

Suplementary Material_Sanders_etal_2022

## Data Availability

Raw imaging data and sensitive information cannot be openly shared due to privacy issues. Non-sensitive data can be made available upon reasonable request to the authors.

## Abbreviations

MRI: Magnetic resonance imaging
MVPA: moderate- to vigorous intensity physical activity
LPA: Low intensive physical activity
CBF: Cerebral blood flow
ASL: Arterial spin labelling

## Acknowledgments

We are very thankful to those who participated in the StrokeMRI study. The study was funded by the Research Council of Norway [249795, 248238, 276082], the South-Eastern Norway Regional Health Authority [2014097, 2015044, 2015073, 2018037, 2018076, 2019107, 2020086], the Norwegian ExtraFoundation for Health and Rehabilitation [2015/ FO5146], the European Research Council under the European Union’s Horizon 2020 research and Innovation program [ERC StG Grant 802998], Sunnaas Rehabilitation Hospital HT, and the Department of Psychology, University of Oslo.

## Notes

### Competing Interest Statement

The authors have declared no competing interest.

### Author Declarations

The Regional Committees for Medical and Health Research Ethics for the South-Eastern Norway gave ethical approval for this study (REK approvals 2014/ 694, 2015/1282).

