## Supplementary material for "Associations between everyday activities and arterial spin labelling-derived cerebral blood flow: A longitudinal study in community-dwelling elderly volunteers": Suplementary Material_Sanders_etal_2022

[a] NORMENT, Division of Mental Health and Addiction, Oslo University Hospital & Institute of Clinical Medicine, University of Oslo, Oslo, Norway

[b] Department of Psychology, University of Oslo, Oslo, Norway

[c] Sunnaas Rehabilitation Hospital HT, Nesodden, Norway

[d] Oslo New University College, Oslo, Norway

[e] Department of Psychiatric Research, Diakonhjemmet Hospital, Oslo, Norway

[f] Section for Preventive Cardiology, Department of Endocrinology, Obesity and Preventive Medicine, Oslo University Hospital, Oslo, Norway.

[g] Department of Physics and Computational Radiology, Div. of Radiology and Nuclear Medicine, Oslo University Hospital, Oslo, Norway

[h] Centre of Research and Education in Forensic Psychiatry, Oslo University Hospital, Oslo, Norway

[i] Faculty of Health Sciences, Oslo Metropolitan University, Norway

[j] Norwegian Directorate of Health, Oslo, Norway

[k] KG Jebsen Center for Neurodevelopmental Disorders, University of Oslo, Oslo, Norway

**Supplementary Table 1.** Item specific factor loadings (cut-off = 0.5) for Frenchay Activities Index (FAI)

| Item | Domestic work | Social activities |
|---|---|---|
| FAI: Meals | 0.79 | |
| FAI: Washing Up | 0.74 | |
| FAI: Washing Clothes | 0.76 | |
| FAI: Light Housework | 0.85 | |
| FAI: Heavy Housework | 0.71 | |
| FAI: Local Shopping | 0.61 | |
| FAI: Social | | 0.52 |
| FAI: Traveling | | 0.68 |

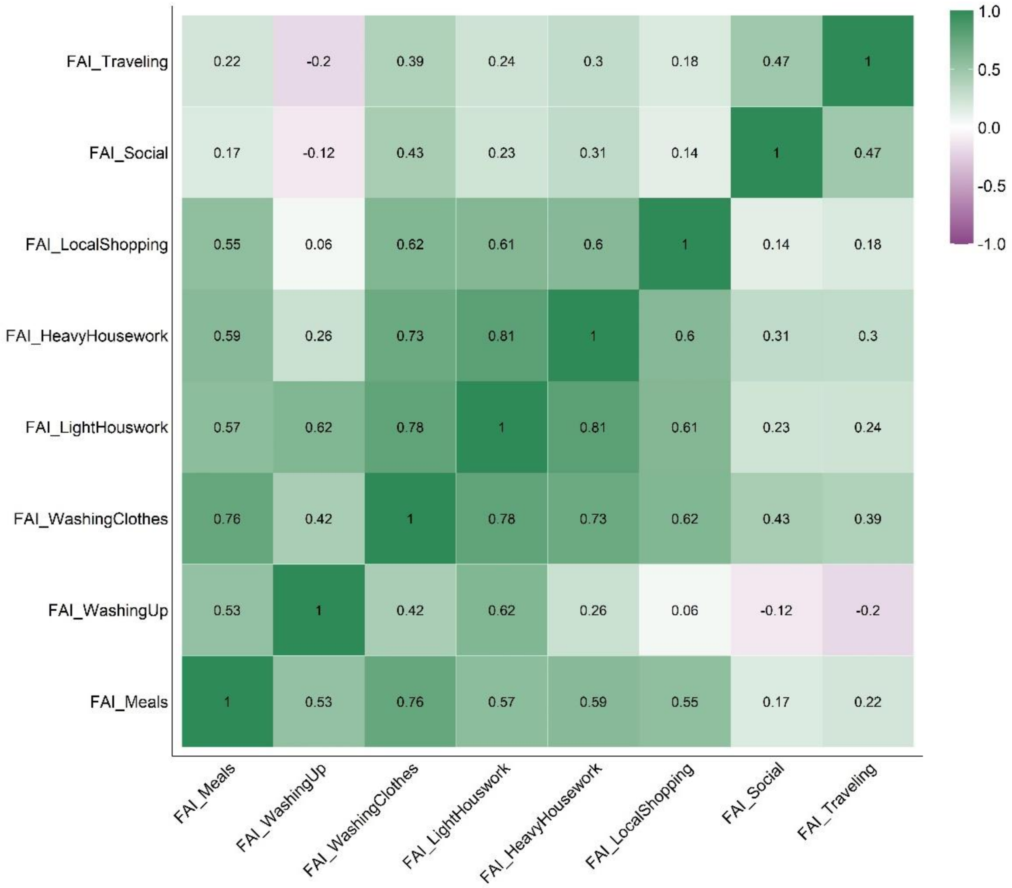


**Supplementary Figure 1.** Pairwise correlation matrix (Pearson´s r) of items from Frenchay Activities index.

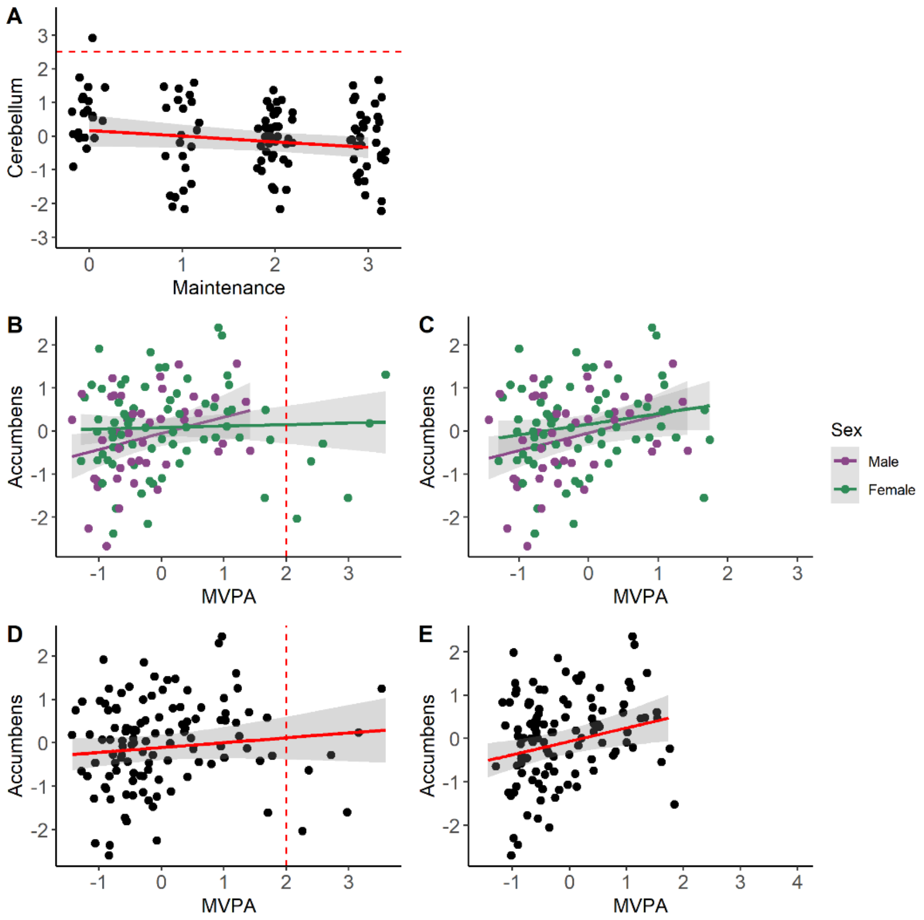


**Supplementary Figure 2.** Examples of results from analysis with and without potentially influential cases. Regression line represent estimated slopes and grey band represents 95 % credible interval (uncertainty). **(A)** Association between cerebellum CBF and maintenance (Frenchey Activities Index). Black points represent observed data, and black point above dashed line denotes influential case with potentially large influence on regression line. (**B)** Interaction effect of sex on the association between accumbens CBF and moderate-to-vigorous physical activity (MVPA). Colored point represents observed data split by sex. Points to the right of the dashed line represents outliers, with higher activity level than two standard deviations above mean, and with potentially large influence on regression line. **(C)** Interaction effect of sex on the association between accumbens CBF and MVPA, after removal of influential cases. **(D)** Results of main analysis of the association between accumbens CBF and MVPA. Black points represent observed data. Points to the right of the dashed line denotes outliers, with higher activity level than two standard deviations above mean, and with potentially large influence on regression line. **(E)** Results of main analysis of the association between accumbens CBF and MVPA, after removal of influential cases.

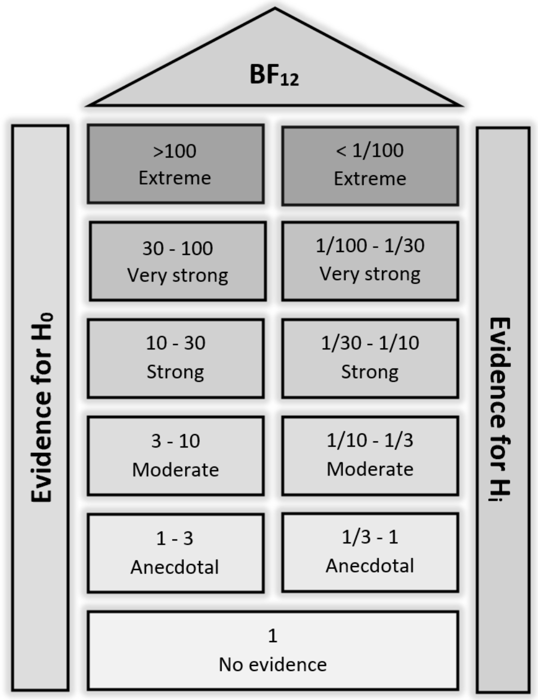


**Supplementary Figure 3.** Evidence categories for the Bayes Factor ($BF_{12}$) (Modified from Jeffreys, 1961).

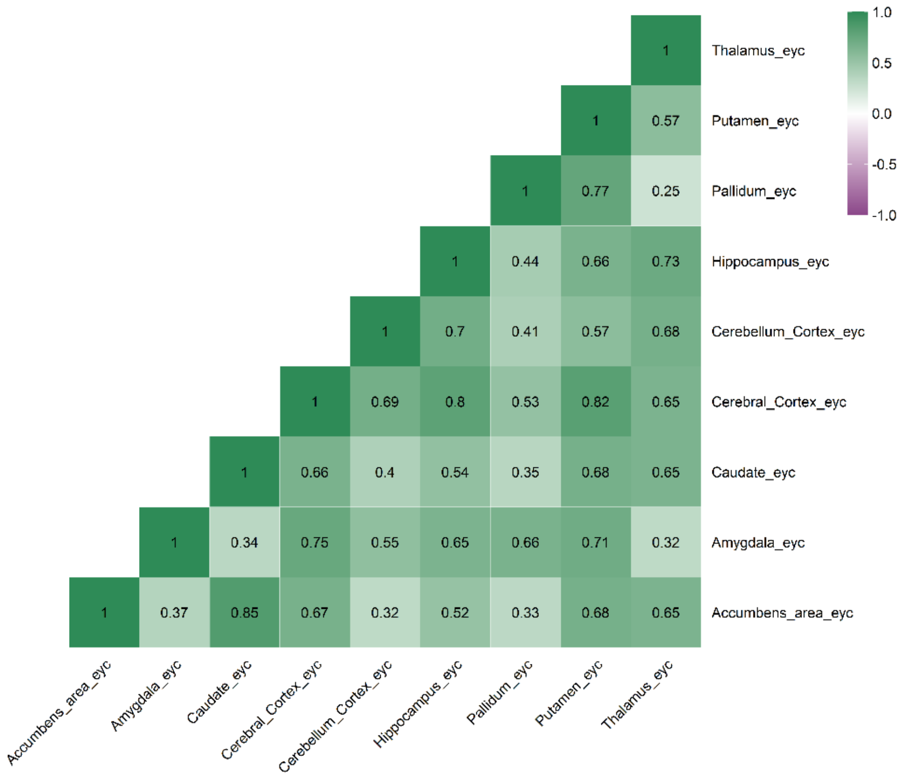


**Supplementary Figure 4.** Pairwise correlation matrix (Pearson´s r) on estimated yearly change in CBF.

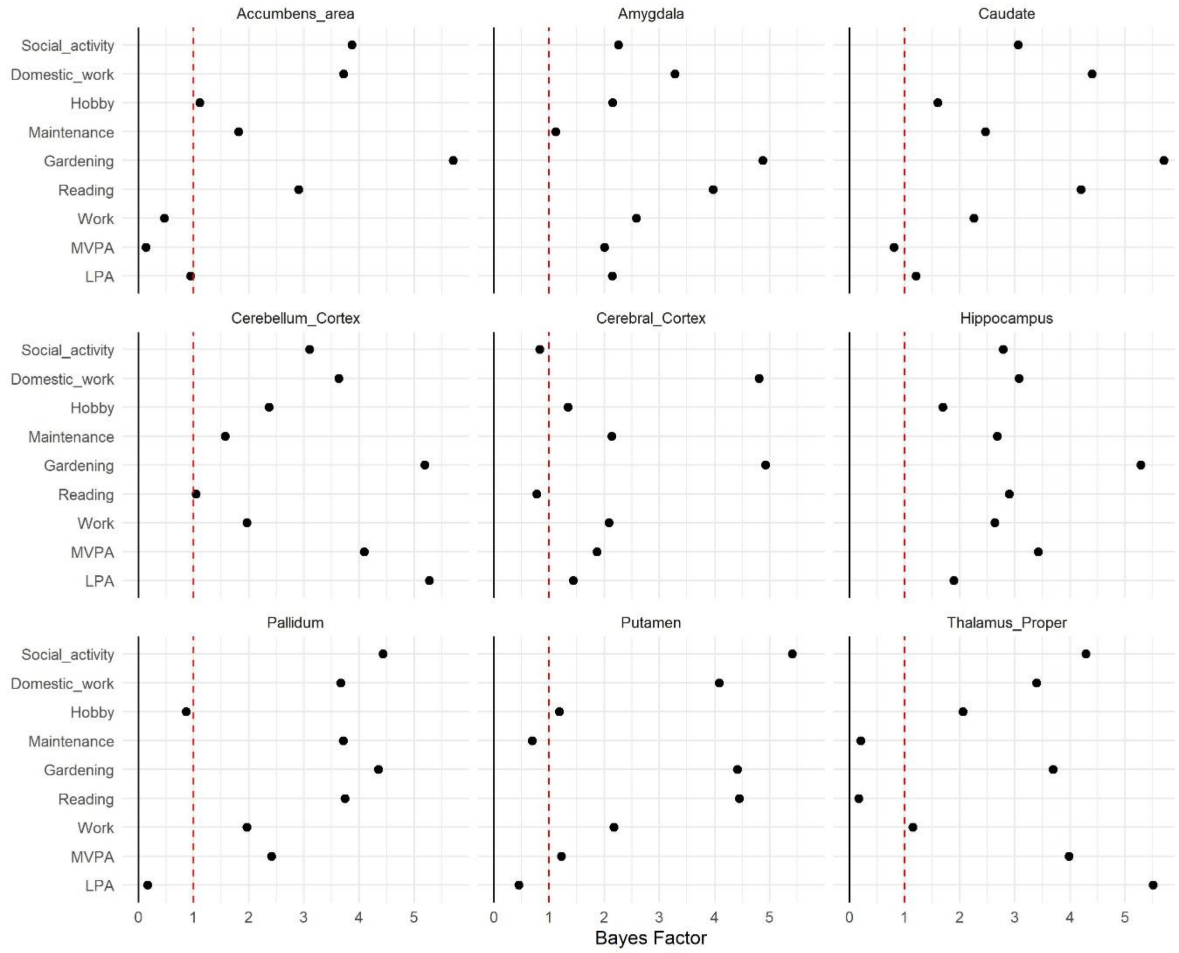


**Supplementary Figure 5.** Bayes Factor, BF12, for the main effect of association between CBF and activity level. Model: CBF ~ activity level + sex + age.

**Supplementary Table 2.** Estimates from the models of the association between CBF and activity level, additionally adjusted for age and sex (*n* = 118)

| Dependent variable: CBF | Activity level | Estimate | Lower95 | Upper95 | Bayes Factor |
|---|---|---|---|---|---|
| **Accumbens area** | LPA | 0.18 | 0.0 | 0.36 | 0.85 |
| | MVPA | 0.31 | 0.08 | 0.53 | 0.13 |
| | Work | -0.35 | -0.71 | 0.03 | 0.47 |
| | Reading | 0.11 | -0.11 | 0.32 | 2.90 |
| | Gardening | -0.02 | -0.2 | 0.15 | 5.71 |
| | Maintenance | -0.14 | -0.32 | 0.04 | 1.82 |
| | Hobby | 0.25 | -0.14 | 0.66 | 1.11 |
| | Domestic_work | -0.07 | -0.27 | 0.15 | 3.72 |
| | Social_activity | 0.08 | -0.1 | 0.27 | 3.87 |
| **Amygdala** | LPA | 0.13 | -0.06 | 0.3 | 2.12 |
| | MVPA | 0.15 | -0.09 | 0.38 | 2.00 |
| | Work | 0.03 | -0.37 | 0.39 | 2.59 |
| | Reading | 0.06 | -0.16 | 0.26 | 3.97 |

| | | | | | |
|---|---|---|---|---|---|
| | Gardening | -0.05 | -0.23 | 0.12 | 4.87 |
| | Maintenance | -0.16 | -0.34 | 0.02 | 1.12 |
| | Hobby | 0.08 | -0.32 | 0.48 | 2.15 |
| | Domestic_work | 0.09 | -0.12 | 0.3 | 3.28 |
| | Social_activity | 0.12 | -0.05 | 0.3 | 2.25 |
| **Caudate** | LPA | 0.16 | -0.02 | 0.34 | 1.19 |
| | MVPA | 0.22 | -0.03 | 0.45 | 0.80 |
| | Work | -0.1 | -0.48 | 0.27 | 2.26 |
| | Reading | 0.05 | -0.17 | 0.25 | 4.20 |
| | Gardening | 0.0 | -0.17 | 0.17 | 5.70 |
| | Maintenance | -0.11 | -0.29 | 0.06 | 2.46 |
| | Hobby | 0.19 | -0.22 | 0.58 | 1.60 |
| | Domestic_work | -0.05 | -0.27 | 0.16 | 4.40 |
| | Social_activity | 0.1 | -0.08 | 0.28 | 3.05 |
| **Cerebellum Cortex** | LPA | 0.02 | -0.15 | 0.2 | 5.60 |
| | MVPA | 0.01 | -0.22 | 0.25 | 4.10 |
| | Work | 0.14 | -0.21 | 0.52 | 1.97 |
| | Reading | 0.18 | -0.03 | 0.38 | 1.04 |
| | Gardening | -0.05 | -0.21 | 0.12 | 5.19 |
| | Maintenance | -0.14 | -0.31 | 0.03 | 1.57 |
| | Hobby | 0.07 | -0.32 | 0.47 | 2.37 |
| | Domestic_work | 0.08 | -0.13 | 0.28 | 3.63 |
| | Social_activity | 0.1 | -0.08 | 0.27 | 3.11 |
| **Cerebral Cortex** | LPA | 0.14 | -0.02 | 0.32 | 1.39 |
| | MVPA | 0.15 | -0.08 | 0.37 | 1.87 |
| | Work | -0.12 | -0.47 | 0.23 | 2.09 |
| | Reading | 0.19 | 0.0 | 0.39 | 0.78 |
| | Gardening | -0.05 | -0.21 | 0.1 | 4.93 |
| | Maintenance | -0.12 | -0.29 | 0.04 | 2.14 |
| | Hobby | 0.22 | -0.14 | 0.59 | 1.34 |
| | Domestic_work | 0.03 | -0.17 | 0.22 | 4.80 |
| | Social_activity | 0.17 | 0.01 | 0.34 | 0.83 |
| **Hippocampus** | LPA | 0.13 | -0.05 | 0.31 | 1.88 |
| | MVPA | 0.07 | -0.16 | 0.31 | 3.42 |
| | Work | -0.06 | -0.42 | 0.32 | 2.63 |
| | Reading | 0.1 | -0.11 | 0.31 | 2.9 |
| | Gardening | -0.04 | -0.21 | 0.13 | 5.28 |
| | Maintenance | -0.11 | -0.28 | 0.07 | 2.68 |
| | Hobby | 0.16 | -0.25 | 0.56 | 1.69 |
| | Domestic_work | 0.1 | -0.12 | 0.3 | 3.07 |

| | | | | | |
|---|---|---|---|---|---|
| | Social_activity | 0.11 | -0.07 | 0.29 | 2.78 |
| **Pallidum** | LPA | 0.24 | 0.05 | 0.41 | 0.22 |
| | MVPA | 0.13 | -0.11 | 0.36 | 2.41 |
| | Work | 0.15 | -0.22 | 0.54 | 1.97 |
| | Reading | -0.07 | -0.28 | 0.15 | 3.74 |
| | Gardening | -0.06 | -0.24 | 0.11 | 4.35 |
| | Maintenance | -0.08 | -0.26 | 0.1 | 3.72 |
| | Hobby | 0.3 | -0.1 | 0.71 | 0.87 |
| | Domestic_work | 0.07 | -0.14 | 0.3 | 3.67 |
| | Social_activity | -0.05 | -0.24 | 0.13 | 4.44 |
| **Putamen** | LPA | 0.2 | 0.02 | 0.38 | 0.49 |
| | MVPA | 0.19 | -0.04 | 0.44 | 1.22 |
| | Work | -0.11 | -0.48 | 0.28 | 2.17 |
| | Reading | -0.02 | -0.23 | 0.2 | 4.45 |
| | Gardening | -0.06 | -0.24 | 0.11 | 4.41 |
| | Maintenance | -0.18 | -0.36 | -0.01 | 0.69 |
| | Hobby | 0.24 | -0.17 | 0.64 | 1.19 |
| | Domestic_work | 0.06 | -0.15 | 0.27 | 4.09 |
| | Social_activity | 0.0 | -0.19 | 0.18 | 5.41 |
| **Thalamus Proper** | LPA | 0.0 | -0.18 | 0.18 | 5.40 |
| | MVPA | 0.03 | -0.2 | 0.26 | 3.98 |
| | Work | -0.24 | -0.62 | 0.12 | 1.15 |
| | Reading | 0.27 | 0.07 | 0.48 | 0.17 |
| | Gardening | -0.08 | -0.25 | 0.09 | 3.69 |
| | Maintenance | -0.23 | -0.4 | -0.06 | 0.20 |
| | Hobby | 0.12 | -0.29 | 0.51 | 2.05 |
| | Domestic_work | -0.09 | -0.3 | 0.12 | 3.40 |
| | Social_activity | 0.06 | -0.12 | 0.24 | 4.29 |

*Note.* LPA: Average daily minutes with low intensity physical activity (< 100 steps per minute), MVPA: Average daily minutes with moderate- to- vigorous physical activity (≥ 100 steps/minute).

**Supplementary Table 3.** Estimates from the models of the main effect of sex[male] on CBF. Model: CBF ~ activity level + sex[male] + age (*n* = 118)

| Dependent variable: CBF | Activity level | Estimate | Lower95 | Upper95 | Bayes Factor |
|---|---|---|---|---|---|
| **Accumbens_area** | LPA | -0.17 | -0.53 | 0.19 | 1.79 |
| | MVPA | -0.23 | -0.59 | 0.12 | 1.20 |
| | Work | -0.13 | -0.5 | 0.23 | 2.09 |
| | Reading | -0.13 | -0.5 | 0.24 | 2.01 |

| | | | | | |
|---|---|---|---|---|---|
| | Gardening | -0.16 | -0.53 | 0.2 | 1.93 |
| | Maintenance | -0.07 | -0.45 | 0.31 | 2.43 |
| | Hobby | -0.15 | -0.51 | 0.21 | 1.99 |
| | Domestic_work | -0.24 | -0.65 | 0.2 | 1.21 |
| | Social_activity | -0.14 | -0.5 | 0.23 | 2.00 |
| **Amygdala** | LPA | -0.37 | -0.74 | -0.02 | 0.39 |
| | MVPA | -0.39 | -0.76 | -0.03 | 0.29 |
| | Work | -0.36 | -0.74 | 0.0 | 0.41 |
| | Reading | -0.37 | -0.73 | 0.0 | 0.43 |
| | Gardening | -0.37 | -0.73 | 0.0 | 0.42 |
| | Maintenance | -0.27 | -0.66 | 0.11 | 1.02 |
| | Hobby | -0.38 | -0.74 | -0.01 | 0.35 |
| | Domestic_work | -0.3 | -0.73 | 0.13 | 0.86 |
| | Social_activity | -0.34 | -0.72 | 0.01 | 0.49 |
| **Caudate** | LPA | -0.23 | -0.58 | 0.14 | 1.16 |
| | MVPA | -0.29 | -0.64 | 0.09 | 0.82 |
| | Work | -0.2 | -0.57 | 0.16 | 1.44 |
| | Reading | -0.21 | -0.58 | 0.16 | 1.46 |
| | Gardening | -0.22 | -0.6 | 0.14 | 1.24 |
| | Maintenance | -0.14 | -0.51 | 0.24 | 2.01 |
| | Hobby | -0.21 | -0.6 | 0.14 | 1.34 |
| | Domestic_work | -0.27 | -0.67 | 0.17 | 1.06 |
| | Social_activity | -0.19 | -0.56 | 0.18 | 1.63 |
| **Cerebellum_Cortex** | LPA | -0.31 | -0.67 | 0.03 | 0.59 |
| | MVPA | -0.29 | -0.65 | 0.07 | 0.81 |
| | Work | -0.28 | -0.63 | 0.09 | 0.85 |
| | Reading | -0.24 | -0.6 | 0.11 | 1.21 |
| | Gardening | -0.28 | -0.63 | 0.09 | 0.86 |
| | Maintenance | -0.19 | -0.56 | 0.18 | 1.52 |
| | Hobby | -0.29 | -0.63 | 0.07 | 0.75 |
| | Domestic_work | -0.21 | -0.62 | 0.2 | 1.40 |
| | Social_activity | -0.26 | -0.62 | 0.09 | 1.00 |
| **Cerebral_Cortex** | LPA | -0.64 | -0.98 | -0.3 | 0.01 |
| | MVPA | -0.65 | -1.01 | -0.31 | 0.00 |
| | Work | -0.61 | -0.95 | -0.26 | 0.01 |
| | Reading | -0.58 | -0.93 | -0.25 | 0.02 |
| | Gardening | -0.61 | -0.95 | -0.27 | 0.01 |
| | Maintenance | -0.55 | -0.91 | -0.19 | 0.05 |
| | Hobby | -0.62 | -0.97 | -0.29 | 0.00 |
| | Domestic_work | -0.6 | -1 | -0.21 | 0.04 |

| | | | | | |
|---|---|---|---|---|---|
| | Social_activity | -0.58 | -0.91 | -0.24 | 0.01 |
| **Hippocampus** | LPA | -0.4 | -0.75 | -0.04 | 0.26 |
| | MVPA | -0.41 | -0.77 | -0.06 | 0.24 |
| | Work | -0.36 | -0.73 | -0.01 | 0.43 |
| | Reading | -0.36 | -0.72 | 0.01 | 0.43 |
| | Gardening | -0.38 | -0.75 | -0.03 | 0.32 |
| | Maintenance | -0.32 | -0.69 | 0.06 | 0.71 |
| | Hobby | -0.39 | -0.74 | -0.02 | 0.32 |
| | Domestic_work | -0.29 | -0.69 | 0.14 | 0.94 |
| | Social_activity | -0.36 | -0.72 | 0 | 0.41 |
| **Pallidum** | LPA | -0.08 | -0.44 | 0.27 | 2.32 |
| | MVPA | -0.15 | -0.51 | 0.2 | 1.97 |
| | Work | -0.12 | -0.49 | 0.25 | 2.11 |
| | Reading | -0.16 | -0.54 | 0.22 | 1.88 |
| | Gardening | -0.12 | -0.48 | 0.26 | 2.15 |
| | Maintenance | -0.08 | -0.46 | 0.31 | 2.28 |
| | Hobby | -0.12 | -0.49 | 0.24 | 2.17 |
| | Domestic_work | -0.07 | -0.49 | 0.37 | 2.13 |
| | Social_activity | -0.16 | -0.53 | 0.22 | 1.89 |
| **Putamen** | LPA | -0.25 | -0.6 | 0.1 | 1.10 |
| | MVPA | -0.31 | -0.68 | 0.05 | 0.64 |
| | Work | -0.23 | -0.6 | 0.13 | 1.21 |
| | Reading | -0.26 | -0.65 | 0.09 | 1.02 |
| | Gardening | -0.23 | -0.59 | 0.16 | 1.32 |
| | Maintenance | -0.12 | -0.51 | 0.25 | 2.11 |
| | Hobby | -0.24 | -0.61 | 0.11 | 1.07 |
| | Domestic_work | -0.19 | -0.61 | 0.24 | 1.52 |
| | Social_activity | -0.25 | -0.63 | 0.11 | 1.09 |
| **Thalamus_Proper** | LPA | -0.47 | -0.83 | -0.1 | 0.11 |
| | MVPA | -0.44 | -0.81 | -0.07 | 0.15 |
| | Work | -0.39 | -0.75 | -0.03 | 0.28 |
| | Reading | -0.34 | -0.7 | 0.01 | 0.48 |
| | Gardening | -0.4 | -0.76 | -0.05 | 0.26 |
| | Maintenance | -0.27 | -0.63 | 0.1 | 0.90 |
| | Hobby | -0.42 | -0.79 | -0.07 | 0.21 |
| | Domestic_work | -0.52 | -0.91 | -0.07 | 0.14 |
| | Social_activity | -0.41 | -0.77 | -0.05 | 0.27 |

*Note*. LPA: Average daily minutes with low intensity physical activity (< 100 steps per minute), MVPA: Average daily minutes with moderate- to- vigorous intensity physical activity (≥ 100 steps/minute).

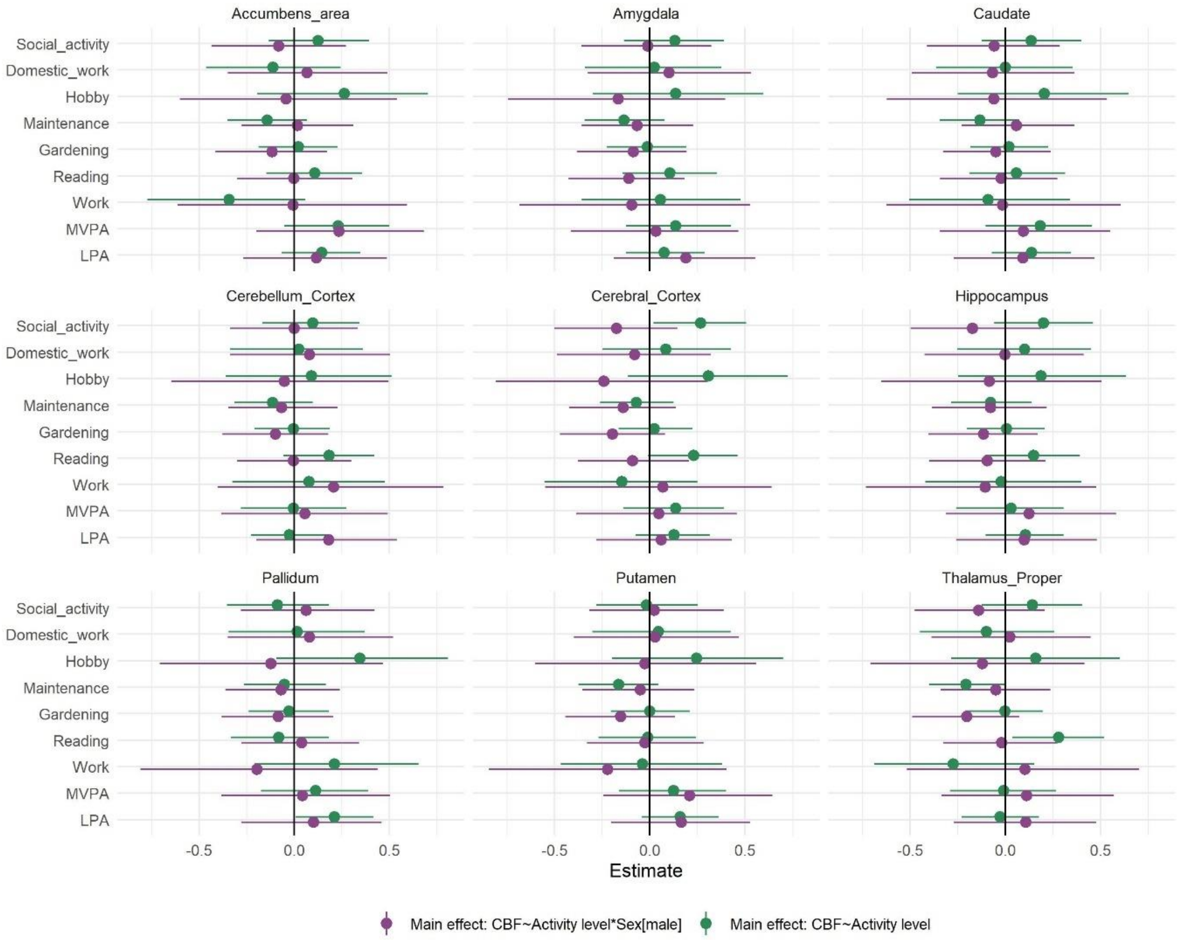


**Supplementary Figure 6.** Parameter estimates reflecting the interaction effect between sex and frequency of activities on CBF (purple) and the association between regional CBF and activity level (green) with 95 % credible interval. Model: CBF ~ activity level × sex + age.

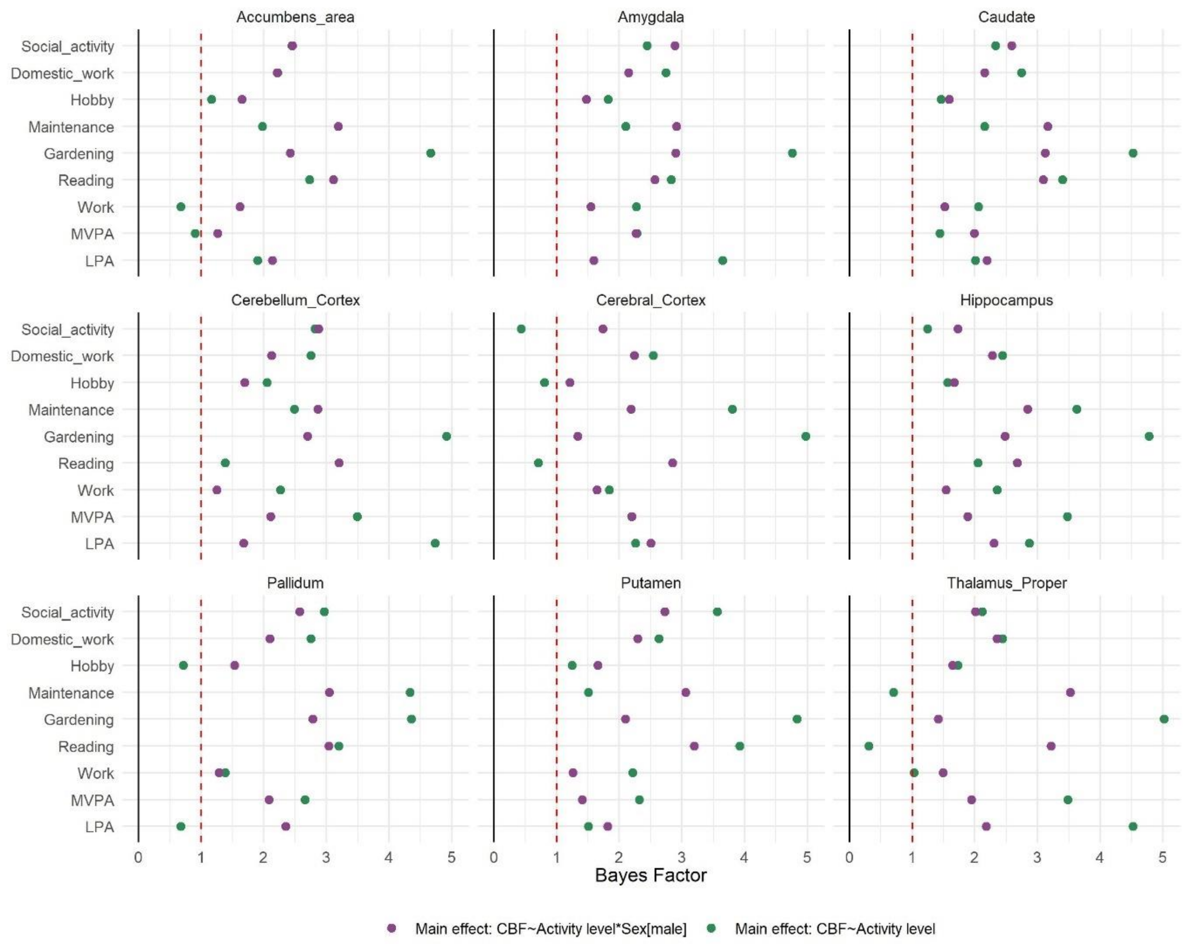


**Supplementary Figure 7.** Bayes Factor, BF12, for the interaction effect between sex and frequency of activities on CBF (purple) and the association between regional CBF and activity level (green). Model: CBF ~ activity level × sex + age

**Supplementary Table 4.** Estimates of the interaction term between activity level and sex on CBF. Model: CBF ~ *activity level* × *sex* + age ($n$ = 118).

| Dependent variable: CBF | Activity_level | Estimate | lower95 | upper95 | Bayes_Factor |
|---|---|---|---|---|---|
| **Accumbens area** | LPA | 0.12 | -0.27 | 0.49 | 2.14 |
| | MVPA | 0.23 | -0.2 | 0.68 | 1.26 |
| | Work | -0.01 | -0.61 | 0.59 | 1.62 |
| | Reading | 0.0 | -0.3 | 0.3 | 3.11 |
| | Gardening | -0.12 | -0.41 | 0.17 | 2.43 |
| | Maintenance | 0.02 | -0.28 | 0.31 | 3.19 |
| | Hobby | -0.04 | -0.6 | 0.54 | 1.65 |
| | Domestic_work | 0.07 | -0.35 | 0.49 | 2.22 |
| | Social_activity | -0.08 | -0.43 | 0.27 | 2.45 |
| **Amygdala** | LPA | 0.19 | -0.19 | 0.55 | 1.59 |
| | MVPA | 0.03 | -0.42 | 0.47 | 2.26 |

| | | | | | |
|---|---|---|---|---|---|
| | Work | -0.09 | -0.68 | 0.53 | 1.54 |
| | Reading | -0.11 | -0.43 | 0.18 | 2.57 |
| | Gardening | -0.09 | -0.38 | 0.19 | 2.90 |
| | Maintenance | -0.07 | -0.36 | 0.23 | 2.91 |
| | Hobby | -0.17 | -0.75 | 0.4 | 1.47 |
| | Domestic_work | 0.1 | -0.33 | 0.53 | 2.15 |
| | Social_activity | -0.01 | -0.36 | 0.32 | 2.89 |
| **Caudate** | LPA | 0.09 | -0.27 | 0.47 | 2.20 |
| | MVPA | 0.1 | -0.35 | 0.55 | 1.99 |
| | Work | -0.02 | -0.63 | 0.6 | 1.52 |
| | Reading | -0.02 | -0.34 | 0.27 | 3.09 |
| | Gardening | -0.05 | -0.33 | 0.24 | 3.12 |
| | Maintenance | 0.06 | -0.23 | 0.36 | 3.16 |
| | Hobby | -0.06 | -0.62 | 0.53 | 1.59 |
| | Domestic_work | -0.07 | -0.49 | 0.36 | 2.15 |
| | Social_activity | -0.06 | -0.41 | 0.29 | 2.59 |
| **Cerebellum_Cortex** | LPA | 0.18 | -0.2 | 0.54 | 1.68 |
| | MVPA | 0.06 | -0.38 | 0.49 | 2.11 |
| | Work | 0.21 | -0.4 | 0.78 | 1.25 |
| | Reading | -0.01 | -0.3 | 0.3 | 3.20 |
| | Gardening | -0.1 | -0.38 | 0.18 | 2.70 |
| | Maintenance | -0.07 | -0.35 | 0.23 | 2.86 |
| | Hobby | -0.05 | -0.65 | 0.49 | 1.70 |
| | Domestic_work | 0.08 | -0.34 | 0.5 | 2.12 |
| | Social_activity | 0.0 | -0.34 | 0.33 | 2.87 |
| **Cerebral_Cortex** | LPA | 0.06 | -0.28 | 0.43 | 2.50 |
| | MVPA | 0.05 | -0.39 | 0.46 | 2.20 |
| | Work | 0.07 | -0.55 | 0.64 | 1.64 |
| | Reading | -0.09 | -0.38 | 0.21 | 2.85 |
| | Gardening | -0.2 | -0.47 | 0.08 | 1.33 |
| | Maintenance | -0.14 | -0.43 | 0.14 | 2.19 |
| | Hobby | -0.24 | -0.81 | 0.3 | 1.21 |
| | Domestic_work | -0.08 | -0.49 | 0.32 | 2.24 |
| | Social_activity | -0.17 | -0.5 | 0.14 | 1.73 |
| **Hippocampus** | LPA | 0.1 | -0.26 | 0.48 | 2.30 |
| | MVPA | 0.12 | -0.31 | 0.58 | 1.89 |
| | Work | -0.11 | -0.73 | 0.48 | 1.54 |
| | Reading | -0.09 | -0.4 | 0.21 | 2.67 |
| | Gardening | -0.11 | -0.41 | 0.17 | 2.48 |
| | Maintenance | -0.08 | -0.39 | 0.22 | 2.84 |

| | | | | | |
|---|---|---|---|---|---|
| | Hobby | -0.08 | -0.65 | 0.51 | 1.67 |
| | Domestic_work | 0.0 | -0.43 | 0.41 | 2.28 |
| | Social_activity | -0.17 | -0.5 | 0.19 | 1.73 |
| **Pallidum** | LPA | 0.1 | -0.28 | 0.46 | 2.35 |
| | MVPA | 0.04 | -0.38 | 0.5 | 2.08 |
| | Work | -0.2 | -0.81 | 0.44 | 1.28 |
| | Reading | 0.04 | -0.28 | 0.34 | 3.04 |
| | Gardening | -0.09 | -0.38 | 0.2 | 2.79 |
| | Maintenance | -0.07 | -0.36 | 0.24 | 3.05 |
| | Hobby | -0.12 | -0.71 | 0.47 | 1.53 |
| | Domestic_work | 0.08 | -0.35 | 0.52 | 2.1 |
| | Social_activity | 0.06 | -0.28 | 0.42 | 2.57 |
| **Putamen** | LPA | 0.16 | -0.2 | 0.53 | 1.82 |
| | MVPA | 0.21 | -0.24 | 0.64 | 1.41 |
| | Work | -0.22 | -0.85 | 0.4 | 1.26 |
| | Reading | -0.03 | -0.33 | 0.28 | 3.19 |
| | Gardening | -0.15 | -0.44 | 0.13 | 2.09 |
| | Maintenance | -0.05 | -0.35 | 0.23 | 3.06 |
| | Hobby | -0.03 | -0.6 | 0.56 | 1.66 |
| | Domestic_work | 0.03 | -0.4 | 0.47 | 2.29 |
| | Social_activity | 0.02 | -0.32 | 0.39 | 2.73 |
| **Thalamus_Proper** | LPA | 0.11 | -0.27 | 0.48 | 2.18 |
| | MVPA | 0.11 | -0.34 | 0.57 | 1.95 |
| | Work | 0.1 | -0.52 | 0.7 | 1.49 |
| | Reading | -0.02 | -0.33 | 0.27 | 3.22 |
| | Gardening | -0.2 | -0.49 | 0.07 | 1.41 |
| | Maintenance | -0.05 | -0.34 | 0.24 | 3.52 |
| | Hobby | -0.12 | -0.71 | 0.42 | 1.64 |
| | Domestic_work | 0.02 | -0.39 | 0.45 | 2.35 |
| | Social_activity | -0.14 | -0.48 | 0.21 | 2.01 |

*Note.* LPA: Average daily minutes with low intensity physical activity (< 100 steps per minute), MVPA: Average daily minutes with moderate- to- vigorous intensity physical activity (≥ 100 steps/minute). All models adjusted for sex and age, with subject as random effect.

**Supplementary Table 5.** Estimates of the main effect of activity level from the models including interaction term between activity level and sex on CBF. Model: CBF ~ *activity level* × sex + age (*n* = 118).

| Dependent variable: CBF | Activity_level | Estimate | lower95 | upper95 | Bayes_Factor |
|---|---|---|---|---|---|
| **Accumbens_area** | LPA | 0.14 | -0.07 | 0.35 | 1.9 |
| | MVPA | 0.23 | -0.05 | 0.5 | 0.9 |

| | | | | | |
|---|---|---|---|---|---|
| | Work | -0.34 | -0.77 | 0.06 | 0.67 |
| | Reading | 0.11 | -0.15 | 0.36 | 2.73 |
| | Gardening | 0.02 | -0.19 | 0.23 | 4.66 |
| | Maintenance | -0.14 | -0.35 | 0.07 | 1.98 |
| | Domestic_work | -0.11 | -0.46 | 0.24 | 2.22 |
| | Social_activity | 0.12 | -0.13 | 0.39 | 2.45 |
| **Amygdala** | LPA | 0.08 | -0.12 | 0.29 | 3.65 |
| | MVPA | 0.14 | -0.13 | 0.43 | 2.28 |
| | Work | 0.06 | -0.36 | 0.48 | 2.27 |
| | Reading | 0.1 | -0.14 | 0.35 | 2.83 |
| | Gardening | -0.01 | -0.23 | 0.19 | 4.76 |
| | Maintenance | -0.14 | -0.34 | 0.08 | 2.1 |
| | Domestic_work | 0.02 | -0.34 | 0.37 | 2.74 |
| | Social_activity | 0.13 | -0.14 | 0.39 | 2.44 |
| **Caudate** | LPA | 0.14 | -0.07 | 0.34 | 2.01 |
| | MVPA | 0.18 | -0.1 | 0.45 | 1.44 |
| | Work | -0.09 | -0.5 | 0.34 | 2.06 |
| | Reading | 0.06 | -0.19 | 0.31 | 3.4 |
| | Gardening | 0.02 | -0.18 | 0.23 | 4.53 |
| | Maintenance | -0.13 | -0.35 | 0.07 | 2.16 |
| | Domestic_work | 0.0 | -0.37 | 0.35 | 2.75 |
| | Social_activity | 0.13 | -0.13 | 0.4 | 2.33 |
| **Cerebellum_Cortex** | LPA | -0.03 | -0.23 | 0.18 | 4.73 |
| | MVPA | 0.0 | -0.28 | 0.27 | 3.49 |
| | Work | 0.08 | -0.32 | 0.48 | 2.27 |
| | Reading | 0.18 | -0.06 | 0.42 | 1.38 |
| | Gardening | -0.01 | -0.21 | 0.19 | 4.91 |
| | Maintenance | -0.11 | -0.31 | 0.1 | 2.49 |
| | Domestic_work | 0.02 | -0.34 | 0.36 | 2.75 |
| | Social_activity | 0.1 | -0.17 | 0.34 | 2.81 |
| **Cerebral_Cortex** | LPA | 0.13 | -0.08 | 0.31 | 2.26 |
| | MVPA | 0.13 | -0.14 | 0.39 | 2.19 |
| | Work | -0.15 | -0.55 | 0.25 | 1.84 |
| | Reading | 0.23 | -0.01 | 0.46 | 0.71 |
| | Gardening | 0.02 | -0.16 | 0.22 | 4.98 |
| | Maintenance | -0.07 | -0.26 | 0.12 | 3.8 |
| | Domestic_work | 0.08 | -0.25 | 0.43 | 2.54 |
| | Social_activity | 0.27 | 0.02 | 0.51 | 0.43 |
| **Hippocampus** | LPA | 0.11 | -0.1 | 0.31 | 2.86 |
| | MVPA | 0.03 | -0.26 | 0.31 | 3.47 |

| | | | | | |
|---|---|---|---|---|---|
| | Work | -0.02 | -0.42 | 0.4 | 2.36 |
| | Reading | 0.15 | -0.11 | 0.39 | 2.05 |
| | Gardening | 0.01 | -0.2 | 0.21 | 4.78 |
| | Maintenance | -0.08 | -0.28 | 0.14 | 3.62 |
| | Domestic_work | 0.1 | -0.25 | 0.45 | 2.44 |
| | Social_activity | 0.2 | -0.06 | 0.46 | 1.25 |
| **Pallidum** | LPA | 0.21 | 0.01 | 0.42 | 0.68 |
| | MVPA | 0.11 | -0.17 | 0.39 | 2.66 |
| | Work | 0.21 | -0.21 | 0.65 | 1.38 |
| | Reading | -0.08 | -0.33 | 0.18 | 3.2 |
| | Gardening | -0.03 | -0.24 | 0.18 | 4.35 |
| | Maintenance | -0.05 | -0.27 | 0.16 | 4.34 |
| | Domestic_work | 0.01 | -0.35 | 0.37 | 2.75 |
| | Social_activity | -0.09 | -0.35 | 0.18 | 2.97 |
| **Putamen** | LPA | 0.16 | -0.04 | 0.36 | 1.5 |
| | MVPA | 0.12 | -0.16 | 0.4 | 2.32 |
| | Work | -0.04 | -0.47 | 0.38 | 2.21 |
| | Reading | -0.01 | -0.27 | 0.24 | 3.92 |
| | Gardening | 0.0 | -0.2 | 0.21 | 4.84 |
| | Maintenance | -0.16 | -0.37 | 0.04 | 1.51 |
| | Domestic_work | 0.04 | -0.3 | 0.42 | 2.63 |
| | Social_activity | -0.02 | -0.28 | 0.25 | 3.56 |
| **Thalamus_Proper** | LPA | -0.03 | -0.23 | 0.18 | 4.52 |
| | MVPA | -0.01 | -0.29 | 0.27 | 3.48 |
| | Work | -0.27 | -0.69 | 0.15 | 1.03 |
| | Reading | 0.28 | 0.04 | 0.52 | 0.31 |
| | Gardening | 0.0 | -0.21 | 0.19 | 5.02 |
| | Maintenance | -0.21 | -0.4 | 0.0 | 0.7 |
| | Domestic_work | -0.1 | -0.45 | 0.25 | 2.44 |
| | Social_activity | 0.14 | -0.12 | 0.4 | 2.11 |

*Note*. LPA: Average daily minutes with low intensity physical activity (< 100 steps per minute), MVPA: Average daily minutes with moderate- to- vigorous intensity physical activity (≥ 100 steps/minute). All models adjusted for sex and age, with subject as random effect.

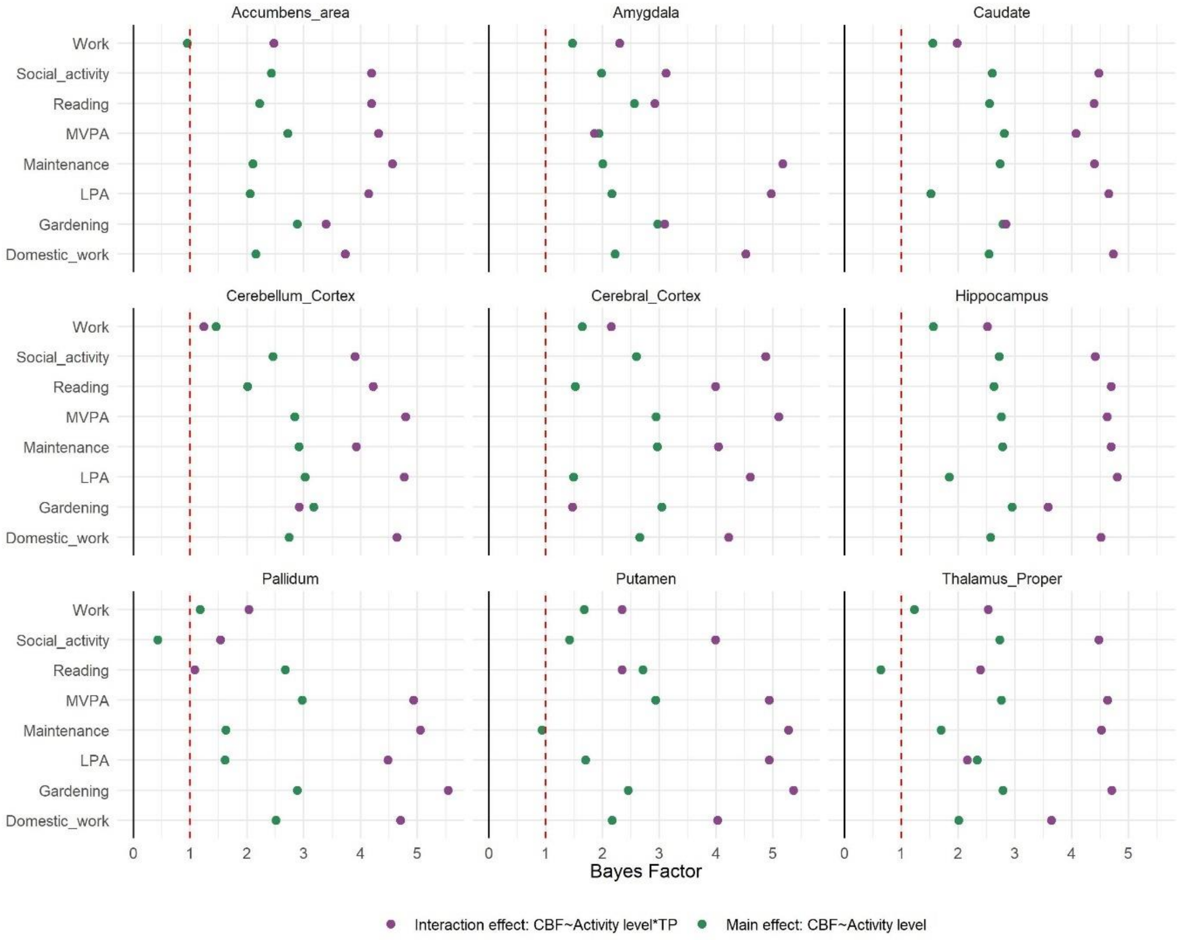


**Supplementary Figure 8.** Bayes Factor, BF12, for the interaction effect between time and frequency of activities on CBF (purple) and the association between regional CBF and activity level (green). Model: CBF~ activity measure × timepoint + age + sex + (1|subject)

**Supplementary Table 6.** Estimates from the models of the interaction effect between time and frequency of activities on CBF. Model: CBF ~ *activity level* × *timepoint* + age +sex + (1|subject) (*n* = 86)

| Dependent variable: CBF | Activity level | Estimate | Lower95 | Upper95 | Bayes Factor |
|---|---|---|---|---|---|
| **Accumbens_area** | LPA | 0.03 | -0.2 | 0.25 | 4.14 |
| | MVPA | 0.04 | -0.19 | 0.27 | 4.31 |
| | Work | -0.02 | -0.42 | 0.38 | 2.47 |
| | Reading | 0.0 | -0.22 | 0.24 | 4.19 |
| | Gardening | -0.09 | -0.3 | 0.11 | 3.39 |
| | Maintenance | -0.04 | -0.25 | 0.17 | 4.56 |
| | Hobby | -0.1 | -0.5 | 0.32 | 2.06 |
| | Domestic_work | 0.07 | -0.16 | 0.3 | 3.73 |
| | Social_activity | 0.0 | -0.24 | 0.23 | 4.19 |
| **Amygdala** | LPA | 0.0 | -0.21 | 0.2 | 4.97 |
| | MVPA | 0.14 | -0.06 | 0.34 | 1.86 |
| | Work | -0.11 | -0.47 | 0.26 | 2.30 |

| | | | | | |
|---|---|---|---|---|---|
| | Reading | -0.1 | -0.31 | 0.11 | 2.92 |
| | Gardening | -0.1 | -0.28 | 0.09 | 3.09 |
| | Maintenance | 0.01 | -0.18 | 0.19 | 5.17 |
| | Domestic_work | -0.03 | -0.23 | 0.18 | 4.52 |
| | Social_activity | 0.1 | -0.1 | 0.31 | 3.11 |
| **Caudate** | LPA | -0.01 | -0.21 | 0.21 | 4.65 |
| | MVPA | 0.06 | -0.15 | 0.27 | 4.07 |
| | Work | -0.13 | -0.5 | 0.25 | 1.98 |
| | Reading | 0.02 | -0.19 | 0.24 | 4.39 |
| | Gardening | -0.11 | -0.29 | 0.09 | 2.84 |
| | Maintenance | -0.06 | -0.25 | 0.14 | 4.4 |
| | Hobby | -0.08 | -0.46 | 0.32 | 2.33 |
| | Domestic_work | 0.01 | -0.2 | 0.23 | 4.73 |
| | Social_activity | 0.02 | -0.2 | 0.23 | 4.48 |
| **Cerebellum_Cortex** | LPA | 0.04 | -0.16 | 0.23 | 4.77 |
| | MVPA | 0.01 | -0.2 | 0.2 | 4.79 |
| | Work | -0.22 | -0.59 | 0.13 | 1.24 |
| | Reading | -0.06 | -0.27 | 0.15 | 4.22 |
| | Gardening | -0.1 | -0.29 | 0.08 | 2.92 |
| | Maintenance | -0.08 | -0.26 | 0.11 | 3.92 |
| | Domestic_work | 0.03 | -0.17 | 0.24 | 4.64 |
| | Social_activity | 0.06 | -0.14 | 0.26 | 3.90 |
| **Cerebral_Cortex** | LPA | -0.05 | -0.24 | 0.14 | 4.60 |
| | MVPA | 0.03 | -0.16 | 0.21 | 5.10 |
| | Work | -0.13 | -0.48 | 0.22 | 2.15 |
| | Reading | -0.06 | -0.26 | 0.14 | 3.99 |
| | Gardening | -0.14 | -0.32 | 0.03 | 1.47 |
| | Maintenance | -0.08 | -0.25 | 0.1 | 4.04 |
| | Domestic_work | -0.06 | -0.25 | 0.14 | 4.22 |
| | Social_activity | -0.04 | -0.23 | 0.14 | 4.87 |
| **Hippocampus** | LPA | 0.01 | -0.2 | 0.22 | 4.80 |
| | MVPA | -0.03 | -0.23 | 0.18 | 4.62 |
| | Work | -0.04 | -0.41 | 0.33 | 2.51 |
| | Reading | 0.0 | -0.22 | 0.21 | 4.69 |
| | Gardening | -0.09 | -0.28 | 0.11 | 3.58 |
| | Maintenance | -0.03 | -0.23 | 0.16 | 4.69 |
| | Domestic_work | 0.03 | -0.18 | 0.24 | 4.51 |
| | Social_activity | 0.04 | -0.18 | 0.26 | 4.41 |
| **Pallidum** | LPA | 0.02 | -0.18 | 0.22 | 4.48 |
| | MVPA | 0.03 | -0.16 | 0.24 | 4.93 |

| | | | | | |
|---|---|---|---|---|---|
| | Work | -0.14 | -0.5 | 0.24 | 2.03 |
| | Reading | -0.19 | -0.39 | 0.02 | 1.08 |
| | Gardening | 0.0 | -0.18 | 0.18 | 5.54 |
| | Maintenance | 0.04 | -0.14 | 0.23 | 5.05 |
| | Hobby | -0.19 | -0.56 | 0.19 | 1.53 |
| | Domestic_work | -0.04 | -0.24 | 0.16 | 4.70 |
| | Social_activity | 0.16 | -0.04 | 0.35 | 1.53 |
| **Putamen** | LPA | 0.03 | -0.17 | 0.23 | 4.93 |
| | MVPA | 0.03 | -0.17 | 0.22 | 4.93 |
| | Work | -0.11 | -0.46 | 0.25 | 2.34 |
| | Reading | -0.13 | -0.32 | 0.08 | 2.34 |
| | Gardening | -0.02 | -0.21 | 0.16 | 5.36 |
| | Maintenance | 0.02 | -0.16 | 0.2 | 5.27 |
| | Hobby | -0.23 | -0.6 | 0.15 | 1.33 |
| | Domestic_work | -0.07 | -0.26 | 0.13 | 4.02 |
| | Social_activity | 0.08 | -0.12 | 0.27 | 3.99 |
| **Thalamus_Proper** | LPA | 0.13 | -0.09 | 0.35 | 2.16 |
| | MVPA | 0.01 | -0.2 | 0.23 | 4.63 |
| | Work | 0.01 | -0.4 | 0.38 | 2.53 |
| | Reading | -0.13 | -0.34 | 0.09 | 2.39 |
| | Gardening | -0.04 | -0.24 | 0.15 | 4.7 |
| | Maintenance | 0.02 | -0.19 | 0.22 | 4.52 |
| | Domestic_work | 0.07 | -0.16 | 0.29 | 3.64 |
| | Social_activity | -0.02 | -0.25 | 0.2 | 4.48 |

*Note.* LPA: Average daily minutes with low intensity physical activity (< 100 steps per minute), MVPA: Average daily minutes with moderate- to- vigorous intensity physical activity (≥ 100 steps/minute). All models adjusted for sex and age, with subject as random effect.

**Supplementary Table 7.** Estimates of the main effect of activity level from the models including interaction term between activity level and time on CBF. Model: CBF ~ *activity level* × timepoint + age + sex+ (1|subject) (*n* = 86).

| Dependent variable: CBF | Activity level | Estimate | Lower95 | Upper95 | Bayes Factor |
|---|---|---|---|---|---|
| **Accumbens_area** | LPA | 0.13 | -0.26 | 0.49 | 2.05 |
| | MVPA | 0.02 | -0.35 | 0.39 | 2.72 |
| | Work | -0.32 | -0.97 | 0.3 | 0.95 |
| | Reading | 0.1 | -0.3 | 0.48 | 2.22 |
| | Gardening | 0.03 | -0.3 | 0.38 | 2.88 |
| | Maintenance | -0.14 | -0.48 | 0.2 | 2.1 |
| | Hobby | 0.1 | -0.53 | 0.76 | 1.4 |

| | | | | | |
|---|---|---|---|---|---|
| | Domestic_work | -0.12 | -0.51 | 0.27 | 2.15 |
| | Social_activity | -0.06 | -0.45 | 0.31 | 2.43 |
| **Amygdala** | LPA | 0.13 | -0.2 | 0.49 | 2.16 |
| | MVPA | -0.15 | -0.5 | 0.18 | 1.94 |
| | Work | 0.14 | -0.47 | 0.75 | 1.47 |
| | Reading | 0.05 | -0.33 | 0.41 | 2.56 |
| | Gardening | 0.01 | -0.31 | 0.33 | 2.97 |
| | Maintenance | -0.15 | -0.47 | 0.18 | 2 |
| | Hobby | 0.07 | -0.57 | 0.7 | 1.53 |
| | Domestic_work | 0.1 | -0.26 | 0.47 | 2.22 |
| | Social_activity | -0.15 | -0.49 | 0.22 | 1.98 |
| **Caudate** | LPA | 0.2 | -0.14 | 0.58 | 1.52 |
| | MVPA | -0.03 | -0.38 | 0.33 | 2.81 |
| | Work | 0 | -0.63 | 0.6 | 1.55 |
| | Reading | 0 | -0.39 | 0.36 | 2.55 |
| | Gardening | 0.07 | -0.26 | 0.4 | 2.79 |
| | Maintenance | -0.08 | -0.41 | 0.25 | 2.74 |
| | Hobby | -0.01 | -0.67 | 0.61 | 1.52 |
| | Domestic_work | -0.05 | -0.41 | 0.33 | 2.54 |
| | Social_activity | -0.07 | -0.42 | 0.31 | 2.6 |
| **Cerebellum_Cortex** | LPA | -0.01 | -0.35 | 0.31 | 3.02 |
| | MVPA | -0.03 | -0.37 | 0.31 | 2.84 |
| | Work | 0.14 | -0.46 | 0.73 | 1.45 |
| | Reading | 0.14 | -0.23 | 0.51 | 2.01 |
| | Gardening | -0.01 | -0.3 | 0.31 | 3.17 |
| | Maintenance | -0.04 | -0.36 | 0.27 | 2.91 |
| | Hobby | 0.25 | -0.36 | 0.87 | 1.15 |
| | Domestic_work | -0.02 | -0.38 | 0.34 | 2.74 |
| | Social_activity | -0.09 | -0.44 | 0.25 | 2.46 |
| **Cerebral_Cortex** | LPA | 0.2 | -0.12 | 0.53 | 1.49 |
| | MVPA | -0.03 | -0.35 | 0.3 | 2.94 |
| | Work | -0.09 | -0.67 | 0.49 | 1.64 |
| | Reading | 0.19 | -0.16 | 0.55 | 1.52 |
| | Gardening | 0.04 | -0.26 | 0.34 | 3.04 |
| | Maintenance | -0.06 | -0.37 | 0.26 | 2.96 |
| | Hobby | 0.25 | -0.38 | 0.85 | 1.19 |
| | Domestic_work | 0.05 | -0.3 | 0.39 | 2.65 |
| | Social_activity | 0.09 | -0.23 | 0.42 | 2.59 |
| **Hippocampus** | LPA | 0.16 | -0.19 | 0.51 | 1.84 |
| | MVPA | 0.02 | -0.31 | 0.38 | 2.76 |

| | | | | | |
|---|---|---|---|---|---|
| | Work | -0.02 | -0.62 | 0.59 | 1.56 |
| | Reading | 0.01 | -0.35 | 0.39 | 2.63 |
| | Gardening | 0.03 | -0.31 | 0.35 | 2.95 |
| | Maintenance | -0.05 | -0.39 | 0.28 | 2.78 |
| | Hobby | 0.07 | -0.56 | 0.72 | 1.5 |
| | Domestic_work | 0.04 | -0.33 | 0.42 | 2.57 |
| | Social_activity | -0.05 | -0.41 | 0.32 | 2.72 |
| **Pallidum** | LPA | 0.19 | -0.16 | 0.53 | 1.61 |
| | MVPA | -0.04 | -0.37 | 0.32 | 2.97 |
| | Work | 0.24 | -0.38 | 0.82 | 1.17 |
| | Reading | 0.03 | -0.33 | 0.4 | 2.67 |
| | Gardening | -0.05 | -0.37 | 0.27 | 2.88 |
| | Maintenance | -0.18 | -0.5 | 0.15 | 1.63 |
| | Hobby | 0.15 | -0.48 | 0.79 | 1.38 |
| | Domestic_work | 0.08 | -0.28 | 0.45 | 2.51 |
| | Social_activity | -0.34 | -0.69 | 0 | 0.43 |
| **Putamen** | LPA | 0.18 | -0.16 | 0.51 | 1.7 |
| | MVPA | -0.01 | -0.36 | 0.32 | 2.93 |
| | Work | 0.03 | -0.59 | 0.62 | 1.68 |
| | Reading | 0.04 | -0.32 | 0.41 | 2.71 |
| | Gardening | -0.11 | -0.42 | 0.22 | 2.45 |
| | Maintenance | -0.25 | -0.56 | 0.08 | 0.93 |
| | Hobby | 0.17 | -0.46 | 0.79 | 1.37 |
| | Domestic_work | 0.13 | -0.21 | 0.5 | 2.17 |
| | Social_activity | -0.21 | -0.54 | 0.14 | 1.42 |
| **Thalamus_Proper** | LPA | -0.1 | -0.45 | 0.29 | 2.33 |
| | MVPA | -0.02 | -0.4 | 0.34 | 2.76 |
| | Work | -0.22 | -0.82 | 0.42 | 1.23 |
| | Reading | 0.32 | -0.05 | 0.7 | 0.64 |
| | Gardening | -0.06 | -0.39 | 0.28 | 2.79 |
| | Maintenance | -0.18 | -0.51 | 0.16 | 1.7 |
| | Hobby | 0.12 | -0.51 | 0.77 | 1.39 |
| | Domestic_work | -0.14 | -0.53 | 0.22 | 2.01 |
| | Social_activity | 0 | -0.36 | 0.38 | 2.73 |

*Note.* LPA: Average daily minutes with low intensity physical activity (< 100 steps per minute), MVPA: Average daily minutes with moderate- to- vigorous intensity physical activity (≥ 100 steps/minute). All models adjusted for sex and age, with subject as random effect.

### References, Supplementary material:

Jeffreys, H. (1961). *Theory of probability*, 3rd edn. Oxford: Oxford University Press.
